# “Urban-Satellite” estimates in the ABCD Study: Linking Neuroimaging and Mental Health to Satellite Imagery Measurements of Macro Environmental Factors

**DOI:** 10.1101/2023.11.06.23298044

**Authors:** Ran Goldblatt, Nathalie Holz, Garrett Tate, Kari Sherman, Selamawit Ghebremicael, Soumitra S Bhuyan, Yazan Al-Ajlouni, Sara Santillanes, Ghermay Araya, Shermaine Abad, Megan M. Herting, Wesley Thompson, Bishal Thapaliya, Ram Sapkota, Jiayuan Xu, Jingyu Liu, the environMENTAL consortium, Gunter Schumann, Vince D. Calhoun

## Abstract

While numerous studies over the last decade have highlighted the important influence of environmental factors on mental health, globally applicable data on physical surroundings are still limited. Access to such data and the possibility to link them to epidemiological studies is critical to unlocking the relationship of environment, brain and behaviour and promoting positive future mental health outcomes. The Adolescent Brain Cognitive Development (ABCD) Study is the largest ongoing longitudinal and observational study exploring brain development and child health among children from 21 sites across the United States. Here we describe the linking of the ABCD study data with satellite-based “Urban-Satellite” (UrbanSat) variables consisting of 11 satellite-data derived environmental indicators associated with each subject’s residential address at their baseline visit, including land cover and land use, nighttime lights, and population characteristics. We present these UrbanSat variables and provide a review of the current literature that links environmental indicators with mental health, as well as key aspects that must be considered when using satellite data for mental health research. We also highlight and discuss significant links of the satellite data variables to the default mode network clustering coefficient and cognition. This comprehensive dataset provides the foundation for large-scale environmental epidemiology research.

## 1. Introduction

The idea of mapping diseases spatially to understand how they relate to the human and physical environment has a rich history of applications, going back to the pioneering work of John Snow, who in 1854 mapped the locations of cholera cases in London to identify its source around a pump at Broad Street [1]. Since then, many studies have highlighted the complex inter-relationships between the physical environment and public health [2,3]. Recent studies emphasize the link between environmental factors and mental disorders [4], with 12-20% of conditions like depression and anxiety attributed to environmental influences [5].

Projected to cause around 250,000 additional deaths annually between 2030 and 2050 [6], climate change also challenges mental health, especially during extreme weather events, with more than two thirds of children experiencing posttraumatic stress symptoms post-disasters (reviewed in [7]) and extreme weather conditions and pandemics elevating anxiety and distress [8,9] [10]. Such detrimental effects on health have spurred the adoption and development of Geographical Information Systems (GIS) technologies, including geospatial and remotely sensed observations for understanding the impacts of epidemics and other health aspects on human’s lives [11].

Newer satellites are enhancing remote sensing for global environmental monitoring [12]. They provide synoptic coverage at various spatial and temporal resolutions [13], allowing understanding of many aspects of Earth’s surface, water, and atmospheric systems, especially in remote areas affected by climate change[14]. As of 2023, 6,718 operational satellites orbit Earth [15], capturing electromagnetic radiation to elucidate land changes and approximate environmental conditions [16], hence allowing tracking of environmental influences on physical and mental health, and supporting disease mapping and epidemiology [16,17]. Despite ever-improving satellite data and more than half of the world’s population living in cities, a gap remains in investigating the impact of the physical environment on mental health.

We introduce “Urban Satellite” variables (UrbanSat) originally developed by Xu et al. [2], measuring population density to represent urbanicity. This refined UrbanSat version, featuring 11 environmental attributes, is part of the Adolescent Brain Cognitive Development℠ Study (ABCD Study®) the largest ongoing U.S. study on child brain development across 21 sites [18] encompassing a cohort of over 11,000 children aged 9-10 with extensive measures on physical and mental health, neurocognition, social and emotional functions, culture, environment, and multimodal brain imaging [19].

In the following sections, we review recent studies on UrbanSat attributes relating to mental health and neuroimaging data and describe our developed variables linking satellite imagery with a USA-wide longitudinal neuroimaging cohort of adolescents.

## 2. Satellite data sources for public health research

The first studies to utilize satellite-based observations for public health applications looked at Aerosol Optical Depth (AOD) data to examine the relation between environmental pollution and autism spectrum disorder (ASD) [20] and impaired adaptive and cognitive functioning [21] following early life exposure to PM_2.5_.

With the launch and availability of satellite data in ever improving spatial, spectral and temporal resolutions, studies have begun examining the relation between other measurable aspects of Earth and public health, such as land cover, land use, landscape structure, vegetation cover, water bodies [22], population distribution and nighttime lights, with applications that include assessing risk areas, mapping diseases [12] and predicting disease distribution [23] in geographical areas that have traditionally been less accessible.

### 2.1. Remote sensing satellites and sensors

Remote spectral imaging began in the 1960s with the Television Infrared Observation Satellite (TIROS) series, initiating experimental weather satellites for systematic Earth imaging [24]. Advancements post-1960s significantly expanded satellite imagery availability, with key satellites marking the evolution (Figure 1).

**Figure 1.**
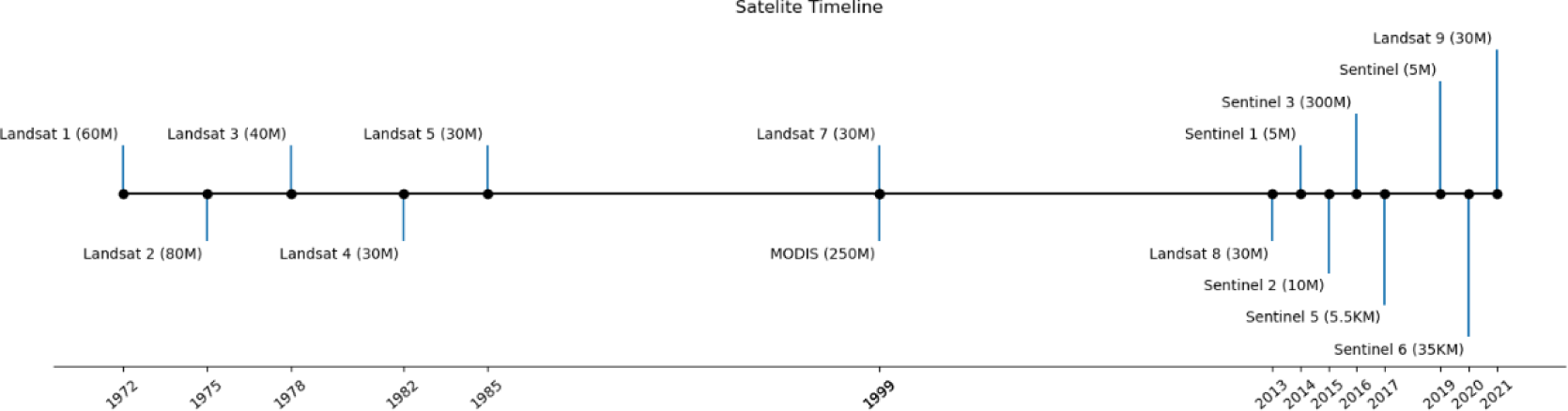
Milestones in the launch of Earth observation satellites, with resolution given in parentheses.

Some of the major satellites used in the field of mental health include Landsat, the Moderate Resolution Imaging Spectroradiometer (MODIS), Sentinel series of satellites, the Defense Meteorological Satellite Program Operational Line Scanner (DMSP/OLS) and high and very-high spatial resolution imagery collected by small satellites and satellite constellations (e.g., RapidEye, Terra Bella and SpaceX) which have become available since the 2000’s.

#### 2.1.1. Landsat

The 1972 launch of Landsat-1, marking the onset of systematic, repetitive Earth observations, revolutionized the accessibility of multispectral satellite imagery and its use across many applications [25]. To date, the Landsat series of satellites has collected more than 10 million satellite images of the Earth. While Landsat data have been used for a wide range of public health applications [26] [27,28], the exploration of land cover and land use impacts on mental health, including schizophrenia [29] and related cognitive facets [30], presents an emerging field of research.

#### 2.1.2. MODIS

The launch of the Moderate Resolution Imaging Spectroradiometer (MODIS) sensor on Terra and Aqua satellites (1999 and 2002, respectively) marked another milestone in satellite records, offering a broad spectral range and field of view, with a spatial resolution of 250 m – 1000 m and 1-2 day revisit periods. Numerous studies have leveraged MODIS data to explore mental health correlations, such as Aerosol Optical Depth (AOD) with schizophrenia [31] and depression [30,32], to evaluate urban greenness’ alleviative effects on depression [33], stratified by age, socioeconomic status, and urbanization [34], and temperature’s relation to depression [35] and cognitive decline [36].

#### 2.1.3. Sentinel

The European Space Agency (ESA) and the European Commission’s Sentinel program, comprises satellites like Sentinel-1 and Sentinel-2, launched in 2014 and 2015 respectively, equipped with radar and multispectral instruments for diverse environmental monitoring. These publicly accessible data have since been applied for public health, including mental health relations to AODs [37], COVID-19 distribution analysis [38] and the impact of the pandemic on winter cropping through management practices [39].

#### 2.1.4. DMSP-OLS and VIIRS

Sensors like the Defense Meteorological Satellite Program Operational Line Scanner (DMSP/OLS) collect data to measure Earth’s nighttime lights (NTL), widely used by the scientific community for various applications [40]. The assumption is that NTL emission, indicative of developed land, can infer the urban extent, economic activity across scales, and light exposure impacts on health [41,42]. DMSP/OLS NTL data has been associated with adverse mental health outcomes, including a higher risk of autism spectrum disorders [43], and increased depressive mood [3]. While a direct link to mental health may not be evident, nighttime light can serve as an important proxy for studying the potential relationships between urbanization, lower socioeconomic status [3], and factors such as disrupted sleep patterns [44] influencing mental health outcomes.

The Visible Infrared Imaging Radiometer Suite (VIIRS) on the Suomi National Polar-orbiting Partnership (S-NPP) satellite, launched in 2011, succeeds DMSP-OLS for low-light Earth imaging. VIIRS day/night band (DNB) offers higher spatial resolution, reduced city center “over-saturation” due to a wider radiometric range and onboard calibration enhancing data quality [45]. These improvements make VIIRS DNB effective for mapping lighting temporal changes [46], offering higher resolution and frequency to better infer socio-economic properties such as population counts, gross regional product (GRP) and electrical power generation [47]. Several studies have relied on VIIRS data for establishing the link between artificial outdoor light and mental disorders, including substance use disorder and depression [48], and sleep disorders [49].

#### 2.1.5. High-resolution satellite EO imagery

Since the 2000’s, there has been a significant advancement in the availability of high-resolution and very high-resolution Electro-Optical (EO) satellite imagery. Very high-resolution data collected by IKONOS, Quickbird and GeoEye, followed by the increasing availability of small satellites and satellite constellations (e.g., RapidEye, Terra Bella and SpaceX) have become available since the 2000’s, allowing access to cheaper and more frequent daily imagery [50]. Several studies employed high to very highresolution imagery in the context of mental health, for instance linking reduced air pollution through green spaces to improved working memory and decreased inattentiveness [51].

We provide in the Supplementary materials a description of methods for satellite data extraction and processing.

## 3. A decade of remotely sensed epidemiology: academic literature trends

Since satellite data’s adoption in public health, studies examining its relation to environmental characteristics and mental health outcomes such as depression and anxiety [52] have steadily increased. Additionally, satellite data has been crucial in responding to public health crises, aiding understanding and response to the COVID-19 pandemic [53–56].

### 3.1. Land Cover and Land Use measurements and their relation to mental health

While discussions surrounding the environmental determinants of physical health are well-established [57–59], the association between Land Use and Land Cover (LULC) patterns and mental disorders has emerged as an increasingly crucial area of research.

A major focus is on urbanization, which has been linked to a higher prevalence of mental health disorders [60], while conversely, the density of green spaces and access to nature within urban environments have shown an inverse relationship with stress levels and the incidence of mental disorders [61]. For example, Dzhambov et al. (2018) [62] used Landsat-8 data to find that neighborhood green spaces moderate the negative impact of traffic noise on mental health.

Evidence suggests that green spaces have a positive influence on development from an early age. For instance, Engemann et al (2019) [61] demonstrated that childhood residence in low-green areas elevated mental illness risk by up to 55% in Danes [61], with both genetic predisposition and green space exposure influencing schizophrenia risk [63]. Similar positive effects on cognitive development [51], partly attributed to a decrease in air pollution levels, and reduced problematic behaviors in children [64] were observed in studies from Barcelona and South Korea, using high-resolution satellite data and the modified soil-adjusted vegetation index (MSAVI) respectively, highlighting the benefits of green environments for children’s mental development. Longitudinal evidence shows short and longterm greenspace exposure near residences reduces adolescent aggressive behaviors, with even slight vegetation increases causing significant behavioral improvement. These associations remained unaffected by sociodemographic and neighborhood quality factors, suggesting greenspace as a preventive measure for urban externalizing problems [65]. Likewise, studies in China and Rome also linked higher vegetation indexes and residential greenness to reduced ADHD symptoms [66] and improved attention-related test performance in children, partly attributed to lowered nitrogen dioxide (NO2) levels [67]. Interestingly, protective effects of green space might be particularly relevant for certain subgroups with children from lower-income households with greenspace access experiencing lower perceived stress against environmental risks like artificial light at night and air pollution [68]. First evidence also points to beneficial effects of green space in the prenatal period. Residential street view-based green space, particularly tree coverage, was associated with lower postpartum depression risk [69], and satellite-based vegetation measurements of green space were linked to a reduced somatization and anxiety symptoms among mothers in a Spanish birth cohort [70]. These results underscore the multifaceted benefits of green environments by highlighting their potential to promote maternal mental well-being during critical developmental periods, ultimately contributing to fostering positive child outcomes.

These benefits extend beyond early development. A comparison of street view and satellite methods assessing green and blue spaces in Beijing revealed an inverse association with geriatric depression [71]. Further, Brown et al., 2018 [72] confirmed the link between green surroundings, measured by NDVI, and mental health in elderly Medicare beneficiaries in Florida, showing 18% and 28% lower risk of Alzheimer’s disease and depression, respectively, in greener areas. Interestingly, as also noted above in children, the positive effect specifically applied to low-income neighborhoods, where an increased greenness was correlated with a 37% lower depression risk compared to wealthier areas suggesting green environments may boost mental well-being in older adults, especially in disadvantaged areas, possibly through promoting physical activity, social interaction, and, thereby, stress reduction. Several studies have established a link between green space and lower stress. For instance, a study on older men from the Caerphilly Prospective Study found urban environment aspects, like housing type and easy street access, associated with reduced psychological distress, highlighting the significance of careful urban planning for healthier communities [73]. Residential tree canopy coverage (TCC) had the potential to counteract the impact of the Covid-19 pandemic on psychological distress with a 1% increase in TCC linked to a 5% decrease in distress prevalence [74].

Evidence shows protective effects of green spaces on mental health globally, yet it is only beginning to be exploredin rapidly urbanizing regions with economic disparities like sub-Saharan Africa. A study from South Africa showcased the role of green environments in mitigating depression, particularly among middle-income individuals and African populations, emphasizing the importance of incorporating environmental considerations into sustainable socioeconomic development efforts in such contexts [75]. While delving into such relationships, it becomes apparent that ecological and economic factors intertwine in distinct ways across countries. As such, urban green space and Gross Domestic Product (GDP) were linked to a nation’s happiness level, with urban green space influencing happiness in wealthier countries and GDP in less wealthy ones. Social support mediated the relationship between urban green space and happiness, while GDP moderated this connection [76].

While numerous studies have highlighted the positive impacts of green spaces on mental health, it is essential to approach this subject with a nuanced perspective taking into account critical mediators of this relationship. A recent Dutch study delved into the long-term relationship between residential greenery exposure and adult suicide mortality, emphasizing individual level risk factors in this association [77]. Likewise, it has been revealed that the presence and severity of affective disorders are associated not just with population density, but with the quality of neighborhood’s socioeconomic, physical, and social characteristics [78]. Further evidence for such indirect pathways was provided by Wang et al. 2020 [79] who found that 62% of the relationship between streetscape greenery and mental wellbeing is mediated by factors like physical activity, stress, air quality, noise, and social cohesion, while NDVI greenery is partially mediated by physical activity and social cohesion, explaining 22% of the association. This suggests that factors beyond urbanization, including elements like socioeconomic status, noise levels, social cohesion, and safety, may significantly influence mental health outcomes.

### 3.3 Remotely sensed nighttime light measurements and mental health

Satellite-measured nighttime lights (NTL) acting as proxies for urbanization, economic and industrial activity, and population distribution, have demonstrated relationships with a variety of mental health outcomes. For instance, Ohayon et al. (2016) [44] relied on DMSP-OLS observations to link higher nighttime lights (NTL) levels with delayed bedtime and wake up time, shorter sleep duration, increased daytime sleepiness, and dissatisfaction with sleep quantity and quality, raising the likelihood of circadian rhythm disorder diagnosis. This relationship was confirmed in a study involving US adolescents associating higher NTL levels with later weeknight bedtimes, shorter sleep durations, and an increased past-year mood and anxiety disorders prevalence [80]. Similarly, in children aged 2 to 18, increased NTL exposure within 500 meters of residence elevated sleep disturbances and sleep disorders risk, particularly among those under 12 [49]. These findings underscore the importance of further research to explore potential interventions for reducing NTL exposure to improve mental and sleep quality.

Important research has highlighted the links between higher NTL and worse mental health outcomes. In South Korea, Min and Min (2018) [42] found significant associations between NTL and depressive symptoms and suicidal behaviors in South Korean adults. Similarly, in the Netherlands, NTL exposure within 100 meters of residence was related to higher depressive symptoms among individuals aged 18 to 65, even after adjusting for confounding factors like air pollution with no such relationship observed for larger 600-meter buffers around residences [81]. This was confirmed by Liao et al. (2022) [3] using data extracted from United Kingdom Biobank Cohort participants to associate higher NTL with increased mental, including depressed mood, tiredness/lethargy, and physical health problems such as obesity as well as more air pollution, less green space, higher economic and neighborhood deprivation and higher household poverty. Leveraging this dataset, a further study established a connection between heightened NTL exposure and an elevated risk of substance use disorder and depression, particularly in individuals with increased iron deposition in the hippocampus and basal ganglia [48]. Several studies have also tied NTL emissions with measurements of people’s perceptions of health and safety, at times showing beneficial effects of NTL, such as feelings of safety and self-reported health [82].

### 3.4 Satellite data and neuroimaging

Despite many opportunities, research exploring the relationship between satellite data, brain features and mental health remains scarce. A seminal stud by Xu et al. [2] provided evidence for a satellite-data derived urbanicity factor being negatively related to medial prefrontal cortex volume and positively to cerebellar vermis volume in Chinese (“CHIMGEN “ sample) and European young adults (“IMAGEN” cohort). Urbanicity also correlated with functional network connectivity, particularly in Chinese participants, and was associated with both positive and negative outcomes, in particular improved social cognition, e.g. perspective-taking but also increased depression symptoms, mediated by brain changes, with susceptibility peaking during mid-childhood and adolescence.

In addition, Dadvand et al. (2018) [83] demonstrated that green neighborhoods may benefit brain development and cognitive function. Specifically, greenness exposure was associated with prefrontal cortex and cerebellar and premotor white matter, predicting improved working memory and reduced inattentiveness.

## 4. UrbanSat variables in the ABCD Study

### 4.1. Sample description

The ABCD Study’s UrbanSat variables consist of 11 key environmental indicators representing land cover characteristics, nighttime lights, population estimates and remote sensing indices in 2017 (see supplemental Figure 1 for a histogram), which were derived from multiple sources, including the Copernicus Global Land Service (CGLS) [84], the Earth Observation Group (EOG) of the Colorado School of Mines [85], WorldPop [86] and Sentinel-2 data processed within Google Earth Engine (GEE) (Table 1). The data are available through the NIMH data archive as part of the ABCD Study 5.0 release (http://dx.doi.org/10.15154/8873-zj65) and include satellite data values linked up to three concurrent addresses for each participant at the baseline study visit when the participants were 9-10 years-old [87], more information is provided in the supplement.

**Table 1.** A description of the 11 key UrbanSat environmental indicators within the ABCD Study.

|  | Description | Units | Source |
| --- | --- | --- | --- |
| 1 | Land Use and Land Cover (LULC) |  |  |
| 1.1 | Percent 2017 Built-up land use | Fraction of total (0 – 1) | CGLS |
| 1.2 | Percent 2017 forest area | Fraction of total (0 – 1) | CGLS |
| 1.3 | Percent 2017 cropland use | Fraction of total (0 – 1) | CGLS |
| 1.4 | Percent 2017 grass area | Fraction of total (0 – 1) | CGLS |
| 1.5 | Percent 2017 permanent inland water area | Fraction of total (0 – 1) | CGLS |
| 1.6 | Percent 2017 seasonal water area | Fraction of total (0 – 1) | CGLS |
| 2 | Nighttime Lights |  |  |
| 2.1 | Total monthly average 2017 night-light radiance | nW/cm<sup>2</sup>/sr | CGLS |
| 3 | Population |  |  |
| 3.1 | Total 2017 population | Number of people | WorldPop |
| 4 | Spectral Indices |  |  |
| 4.1 | Percent 2017 area with NDVI index over 0.2 | Fraction of total (0 – 1) | Sentinel-2 (GEE) |
| 4.2 | Percent 2017 area with NDWI index over 0.3 | Fraction of total (0 – 1) | Sentinel-2 (GEE) |
| 4.3 | Average 2017 NDBI index value | NDBI index value | Sentinel-2 (GEE) |

*Abbreviations: Normalized Difference Vegetation Index (NDVI), Normalized Difference Water Index (NDWI), and Normalized Difference Built-up Index (NDBI), Copernicus Global Land Service (CGLS), Google Earth Engine (GEE)*

### 4.2. Data analysis

Various Urban-Satellite data sources were unified into new raster files with identical parameters (extent, pixel size, and pixel locations) using a custom Python script, aggregated at approximately 1km grid covering the 48 contiguous US states, with detailed methodology available in the supplement.

Input data for each dataset were obtained for the year 2017 to align with the baseline ABCD Study visit timing (October 2016 thru October 2018) and comprised LULC (Figure 2 and Table 2), NTL and population data (Figure 3) and spectral indices (Figure 4), which are described in more detail in the supplement.

**Figure 2.**
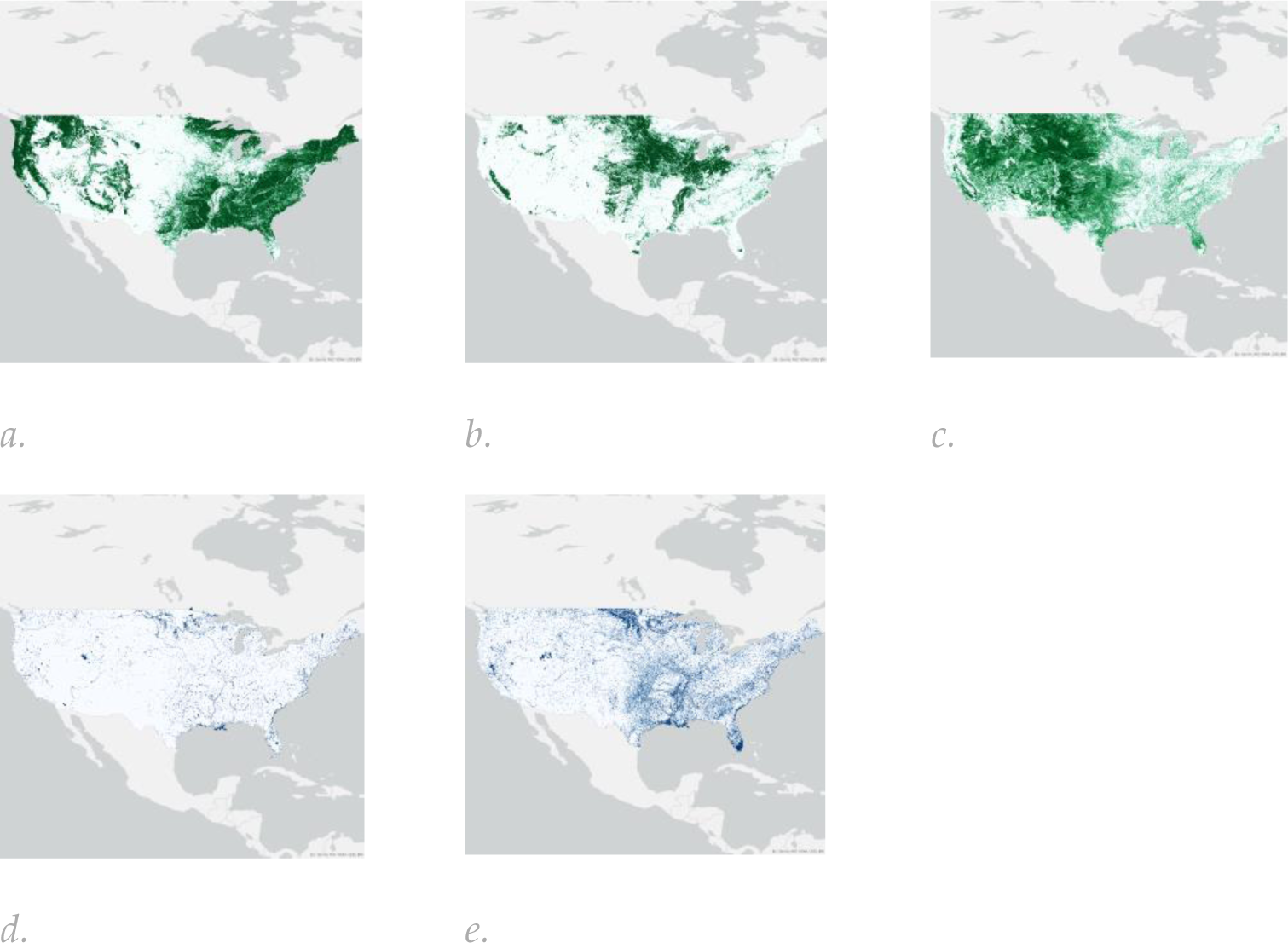
The spatial distribution and characteristics of five of the Land Use / Land Cover (LULC) maps covering the 48 contiguous US states incorporated in the “Urban Satellite” indicators: (a) forest percent; (b) crop percent; (c) grass percent; (d) permanent inland water percent; (e) seasonal water percent.

**Figure 3.**
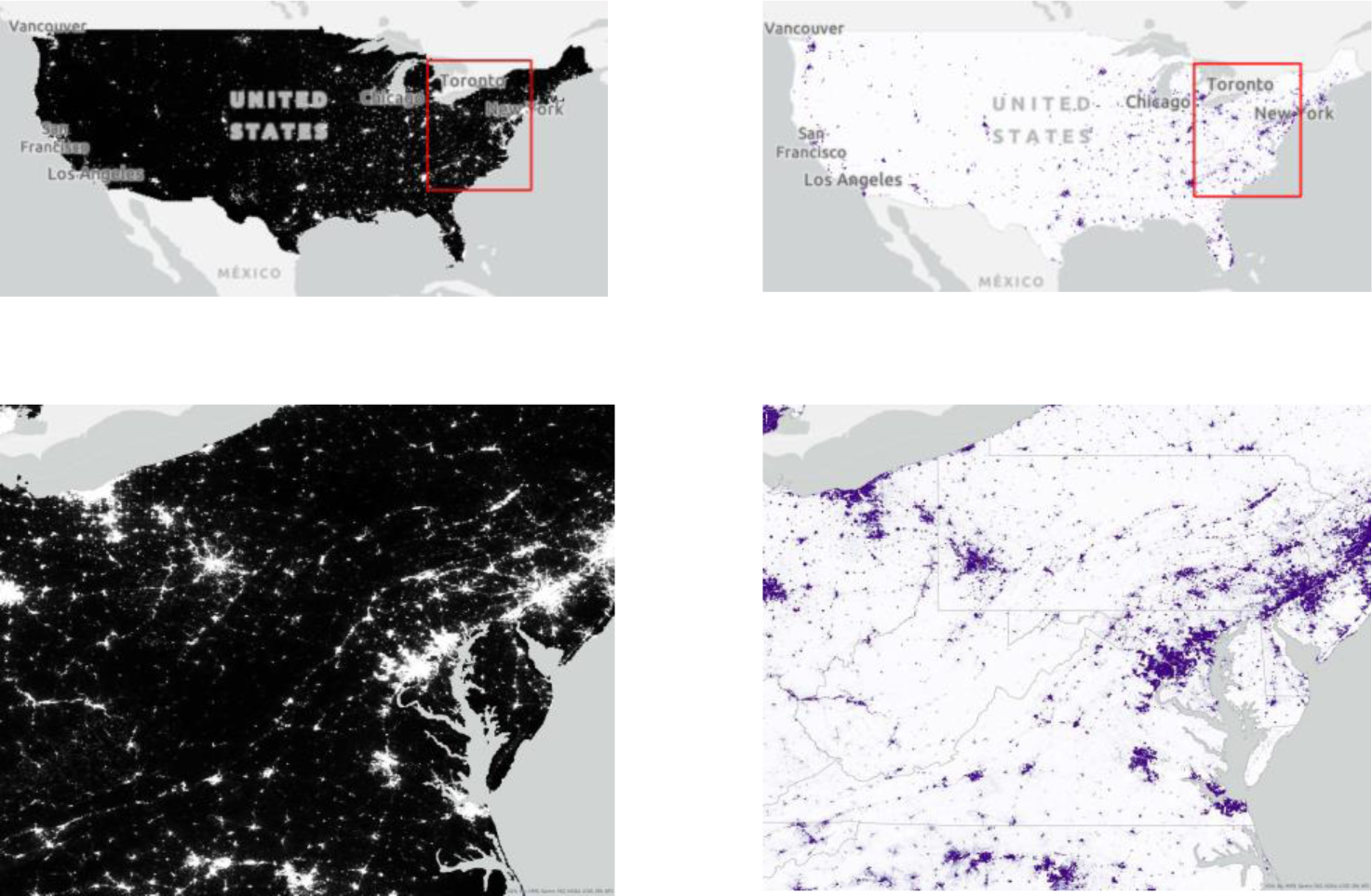
The spatial distribution of (a) nighttime light sourced from the Earth Observation Group (EOG) Annual VNL V2 product [88]. These data provide an average monthly radiance at an original resolution of 15 arc-seconds (approximately 500 m). The VNL 2 data are based on VIIRS satellite observations and include filtering for clouds, removal of fires, and background isolation. Our aggregated nighttime light product provides the sum of annual nighttime light radiance values within each 1 km output pixel. (b) population data from 2017 are based on WorldPop Population Counts [89], specifically the US unconstrained top-down 100 m resolution dataset. These data take population census counts and use other geospatial data to disaggregate census tract information into 100 m by 100 m pixels.

**Figure 4.**
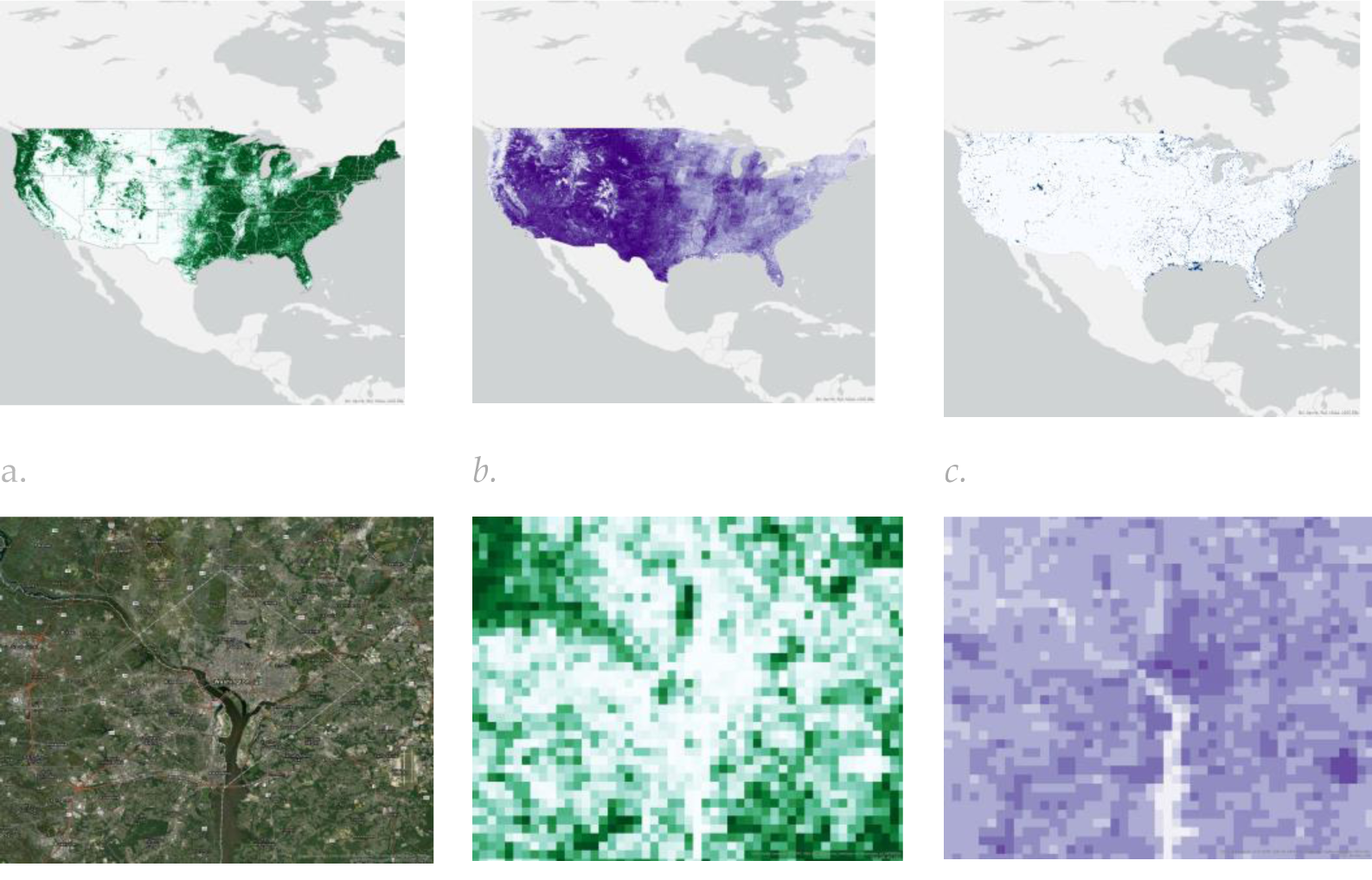
The spatial distribution of (a) Normalized Difference Vegetation Index (NDVI), (b) Normalized Difference Built-up Index (NDBI) and (c) Normalized Difference Water Index (NDWI) within the Urban-Satellite dataset calculated using 2017 Sentinel-2 Multispectral Instrument Level-1C data accessed through Google Earth Engine (GEE); A comparison between the percentage of (e) forest cover and (f) NDBI within the Washington DC area (reference is provided in figure (d)).

**Table 2.**
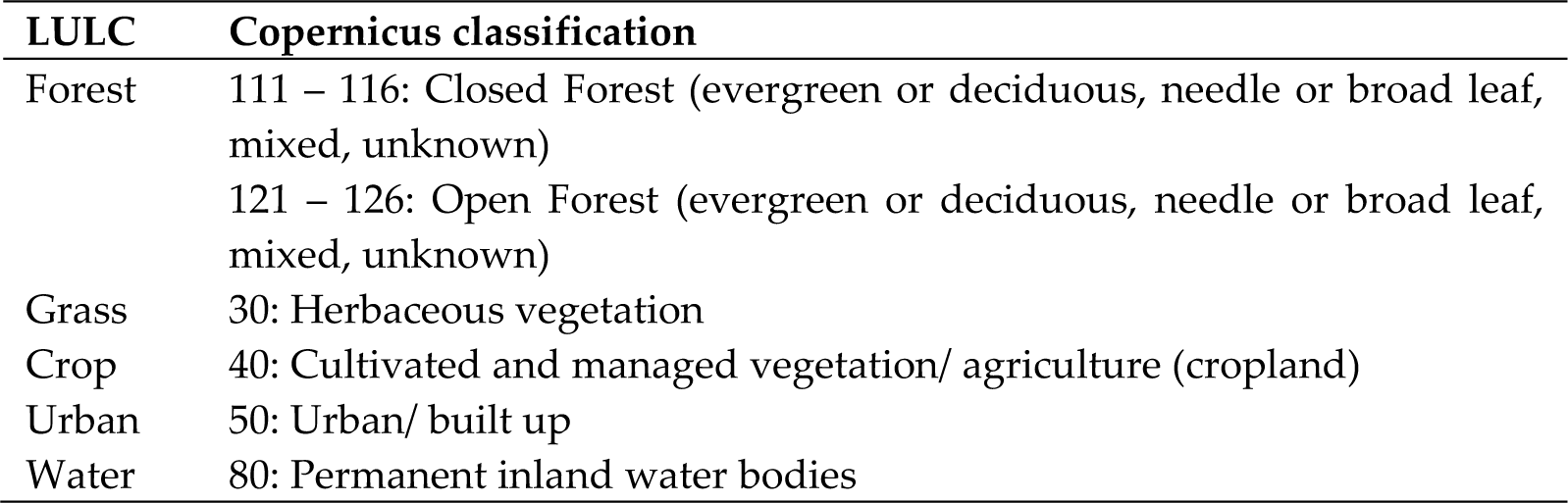
A description of the UrbanSat Copernicus classifications incorporated into the ABCD Study.

| **LULC** | **Copernicus classification** |
| --- | --- |
| Forest | 111 – 116: Closed Forest (evergreen or deciduous, needle or broad leaf, mixed, unknown)<br>121 – 126: Open Forest (evergreen or deciduous, needle or broad leaf, mixed, unknown) |
| Grass | 30: Herbaceous vegetation |
| Crop | 40: Cultivated and managed vegetation/ agriculture (cropland) |
| Urban | 50: Urban/ built up |
| Water | 80: Permanent inland water bodies |

### 4.3. UrbanSat characterization and association with behavioral, cognition and brain function in the ABCD Study

The UrbanSat data in ABCD release 5.0 encompasses 11 variables across 3 baseline addresses, reflecting diverse regional environmental aspects (see supplement and Supplemental Figure 1). A strong correlation emerged between forest and built-up land cover, NDVI, NDBI, nighttime lights, and population, whereas NDWI showed moderate correlations with other indicators (Figure 5, left) (details are available in the supplement).

**Figure 5.**
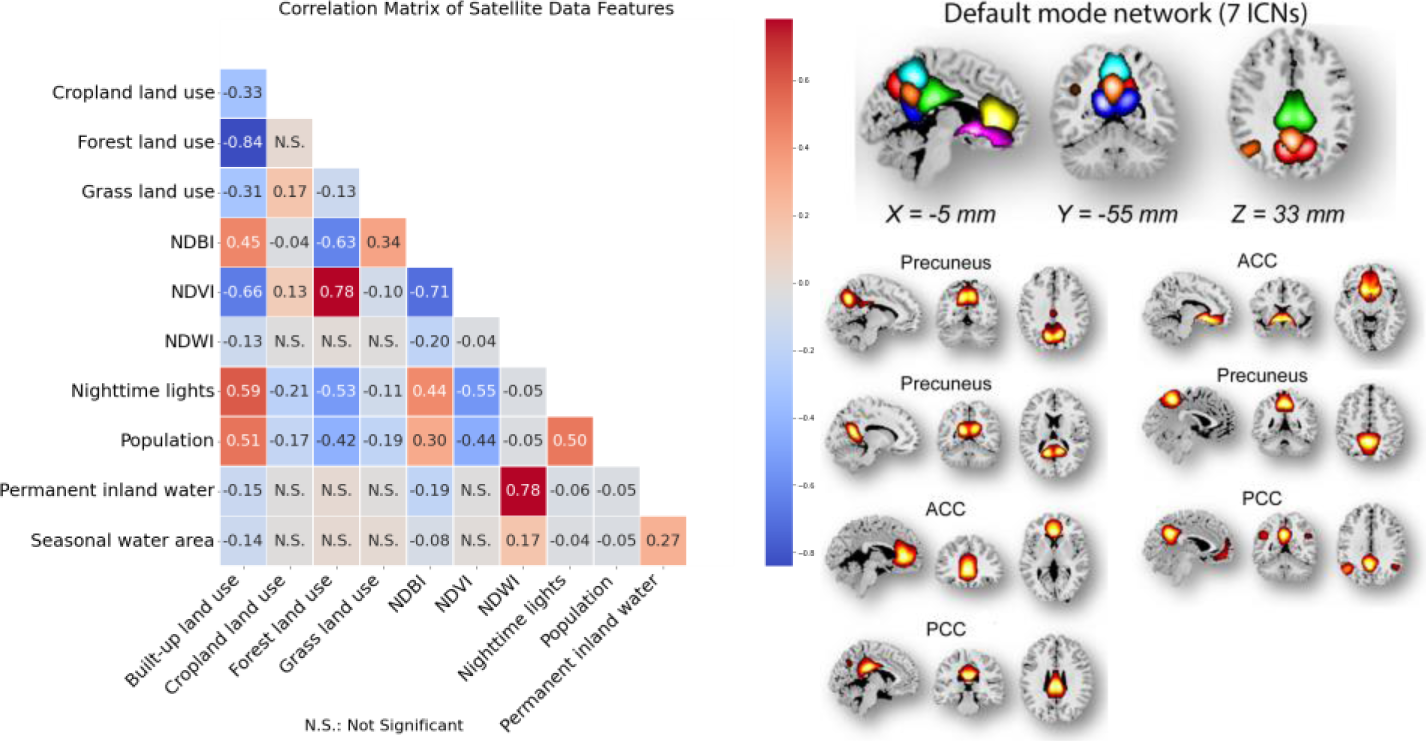
Left: Cross correlation among 11 UrbanSat indicators in the ABCD Study. Right: Seven ICNs within the default mode network. ACC: anterior cingulate cortex, PCC: posterior cingulate cortex.

#### 4.3.1 Behavior and cognition

To demonstrate the influence of UrbanSat indicators on behavior, cognition and brain function, we examined associations with measures from the ABCD Study’s baseline assessments at ages 9-10 and utilized the total problem count from the Child Behavior Checklist (CBCL) [90] and the total score composite from the NIH Toolbox® cognition battery [91] for our analyses. Detailed assessment methodologies and findings are available in the supplement.

#### 4.3.2. Resting state functional MRI data

We extracted 53 intrinsic connectivity networks (ICNs) via a spatially constrained independent component analysis framework, organizing them into seven functional domains (see supplemental Figure 2 and Table S1). We computed Functional Network Connectivity (FNC) and represented the brain as a connected graph, focusing on the default mode network (DMN, Figure 5 right). We calculated the average clustering coefficient of DMN ICNs to represent brain function in our UrbanSat association analyses. Detailed methodologies and equations are provided in the supplementary materials.

#### 4.3.3. Results

We evaluated the correlation between SES (household income and parental education) and UrbanSat indicators (supplementary materials and Table S2). The level of parental education was significantly and negatively correlated to built-up land, NDBI, nighttime lights, and population, and positively correlated to crop land, forest land and NDVI. Household income presented very similar associations with UrbanSat indicators and was most significantly correlated with NDBI. Therefore, due to multi-collinearity two sets of linear mixed effect models were examined with and without SES covariates, and were implemented for UrbanSat association analyses.

Without including SES, UrbanSat indicators were associated with cognition and DMN clustering (except for forest land), with NTL also being associated with problem behavior (Table 3 top panel). Under consideration of SES, NDBI was significantly associated with the cognitive total score, and NTL was significantly associated with DMN clustering coefficient and associated with cognitive score with a trend toward significance (Table 3 bottom panel).

**Table 3.**
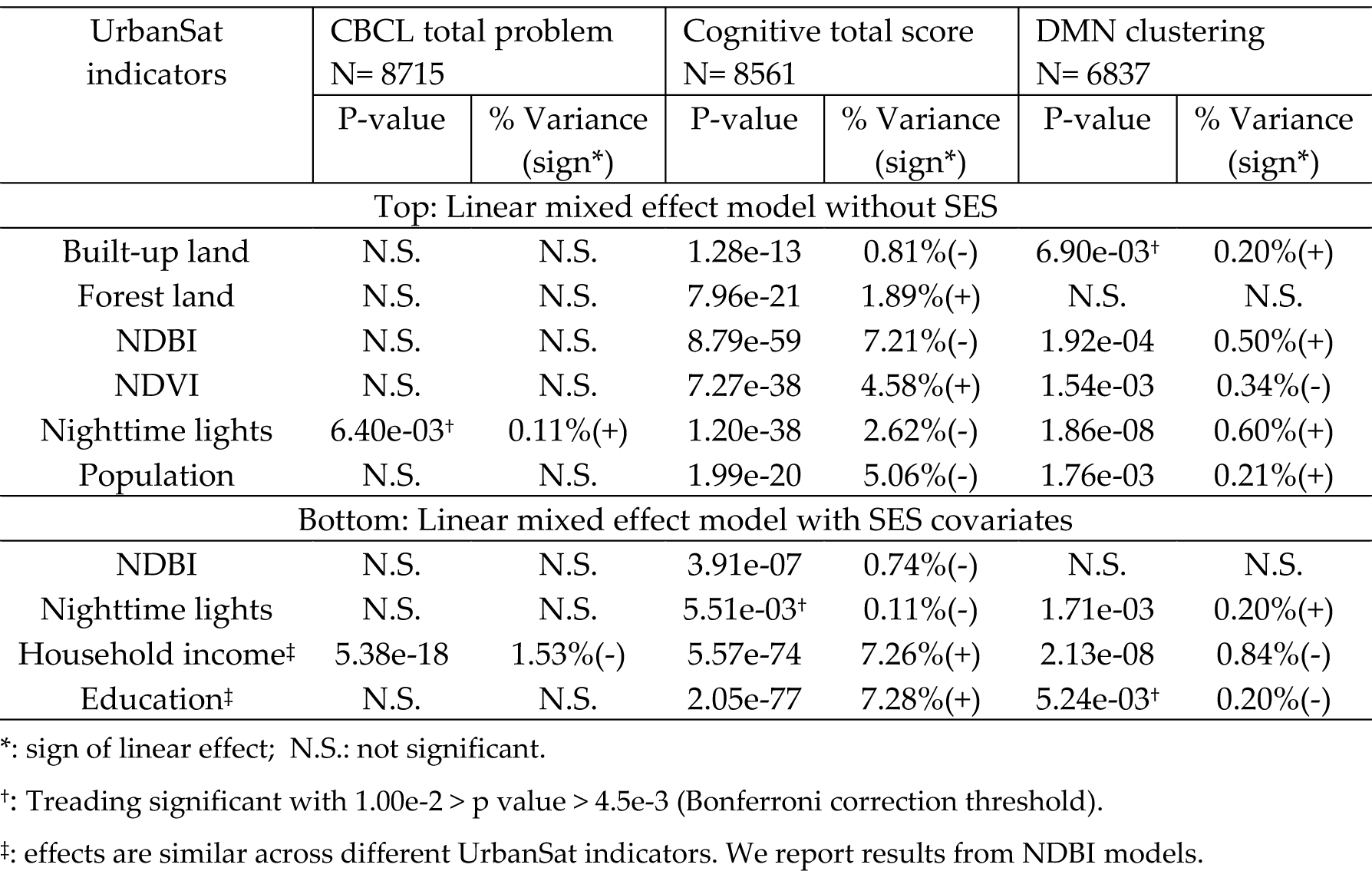
Significant associations of UrbanSat indicators with the CBCL total problem, cognitive total score and DMN clustering coefficient in children ages 9-10 years.

| UrbanSat indicators | CBCL total problem<br>N= 8715 |  | Cognitive total score<br>N= 8561 |  | DMN clustering<br>N= 6837 |  |
| --- | --- | --- | --- | --- | --- | --- |
|  | P-value | % Variance<br>(sign*) | P-value | % Variance<br>(sign*) | P-value | % Variance<br>(sign*) |
| Top: Linear mixed effect model without SES |  |  |  |  |  |  |
| Built-up land | N.S. | N.S. | 1.28e-13 | 0.81%(-) | 6.90e-03<sup>†</sup> | 0.20%(+) |
| Forest land | N.S. | N.S. | 7.96e-21 | 1.89%(+) | N.S. | N.S. |
| NDBI | N.S. | N.S. | 8.79e-59 | 7.21%(-) | 1.92e-04 | 0.50%(+) |
| NDVI | N.S. | N.S. | 7.27e-38 | 4.58%(+) | 1.54e-03 | 0.34%(-) |
| Nighttime lights | 6.40e-03<sup>†</sup> | 0.11%(+) | 1.20e-38 | 2.62%(-) | 1.86e-08 | 0.60%(+) |
| Population | N.S. | N.S. | 1.99e-20 | 5.06%(-) | 1.76e-03 | 0.21%(+) |
| Bottom: Linear mixed effect model with SES covariates |  |  |  |  |  |  |
| NDBI | N.S. | N.S. | 3.91e-07 | 0.74%(-) | N.S. | N.S. |
| Nighttime lights | N.S. | N.S. | 5.51e-03<sup>†</sup> | 0.11%(-) | 1.71e-03 | 0.20%(+) |
| Household income<sup>‡</sup> | 5.38e-18 | 1.53%(-) | 5.57e-74 | 7.26%(+) | 2.13e-08 | 0.84%(-) |
| Education<sup>‡</sup> | N.S. | N.S. | 2.05e-77 | 7.28%(+) | 5.24e-03<sup>†</sup> | 0.20%(-) |

\*: sign of linear effect; N.S.: not significant.
<sup>†</sup>: Trending significant with $1.00e-2 > p \text{ value} > 4.5e-3$ (Bonferroni correction threshold).
<sup>‡</sup>: effects are similar across different UrbanSat indicators. We report results from NDBI models.

## 5. Discussion

In the last few decades understanding the complex links between physical environment and mental health has advanced significantly due to enhanced satellite and airborne sensor technologies. This paper highlights studies exploring mental health correlations with environmental properties captured via satellite imagery, like land cover, urbanization, and NTL. These satellites offer insights into various spatial, spectral and temporal dimensions of human’s physical environment, even in traditionally inaccessible areas.

However, while there is an exponential increase in the availability of satellite records of Earth, integrating them with mental health presents challenges, due to scarce public health datasets on brain and behavior. The ABCD database, with its deep phenotyping information encompassing mental health, cognition, and other health indicators, will aid in disentangling these effects. It captures over 11,800 children with biennial brain scans, and it is considered the largest ongoing study on brain development and child health across 21 US sites.

Within the large domain of the linked external data within the ABCD Study [87], this paper introduces the set of “Urban Satellite” variables, and provides opportunity to understand the interrelation of macro scale environmental factors when the children are 9-10 year-olds with brain development and health.

As proof of concept our simple analyses lend support for the interrelation of environmental factors derived from satellite image with brain and cognitive development, and mental health, while also hinting at the need for careful modeling multicollinearity between UrbanSat indicators and SES indicators. Thereby, we provided evidence for NTL being linked to more dense clustering of DMN with the rest of brain, and NTL and NDBI negatively affecting cognitive ability when controlled for SES. However, the results presented here need to be understood with consideration of limitations. Indeed, the mechanisms linking environmental factors such as UrbanSat variables and mental health, and neurobiological correlates, remain unclear. These connections involve complex physiological, psychological, and social pathways, providing important avenues for future research. For instance, in terms of biologically plausible pathways, the strengthening of physiological systems, such as respiratory health and immune function may act as crucial players linking green space and less build-up to mental health [92]. Further, it has been observed that environmental pollutants, especially fine particles, can breach the protective barrier around the brain, potentially causing damage to the nervous system by triggering neuro-inflammation, disrupting neural signaling, and provoking immune responses [93]. Regarding indirect effects, nature exposure can enhance psychological aspects by reducing negative emotions, while promoting positive feeling [94] and replenishing cognitive resources [95], while also contributing to adaptive perceptions of stressors and the development of self-esteem and new competencies [96]. Moreover, it has been suggested that neighborhood socioeconomic and social aspects, such as diminished social cohesion and reduced safety [78], along with physical activity [79] may mediate the relationship between urbanization and mental health. On the other hand, shared experiences in nature could potentially yield social benefits by encouraging communication, providing support, and fostering cooperation [97]. We expect to see more in-depth investigations of such intricate relationships in the future by linking the UrbanSat indicators with the ABCD data from National Institute of Mental Health’s Data Archive (NDA) (https://dx.doi.org/10.15154/8873-zj65).

## Data Availability

The ABCD data used in this report came from https://dx.doi.org/10.15154/8873-zj65. The data are available through the NIMH data archive as part of the ABCD Study 5.0 release.

## Acknowledgements

Data used in the preparation of this article were obtained from the Adolescent Brain Cognitive Development (ABCD) Study (https://abcdstudy.org), held in the NIMH Data Archive (NDA). This is a multisite, longitudinal study designed to recruit more than 10,000 children aged 9-10 and follow them over 10 years into early adulthood. The ABCD Study is supported by the National Institutes of Health Grants [U01DA041022, U01DA041028, U01DA041048, U01DA041089, U01DA041106, U01DA041117, U01DA041120, U01DA041134, U01DA041148, U01DA041156, U01DA041174, U24DA041123, U24DA041147]. A full list of supporters is available at https://abcdstudy.org/nih-collaborators. A listing of participating sites and a complete listing of the study investigators can be found at https://abcdstudy.org/principal-investigators.html. ABCD consortium investigators designed and implemented the study and/or provided data but did not necessarily participate in analysis or writing of this report. This manuscript reflects the views of the authors and may not reflect the opinions or views of the NIH or ABCD consortium investigators.

The ABCD data repository grows and changes over time. The ABCD data used in this report came from https://dx.doi.org/10.15154/8873-zj65.

Additional support for this work was made possible from grants R01ES032295, R01ES031074, R01DA049238, and NSF grant 2112455, as well as the NSFC grant 82150710554 and Horizon Europe project ‘environMENTAL’ 101057429. Funded by the European Union. Complementary funding was received by UK Research and Innovation (UKRI) under the UK government’s Horizon Europe funding guarantee (10041392 and 10038599). Views and opinions expressed are however those of the author(s) only and do not necessarily reflect those of the European Union or European Health and Digital Executive Agency (HADEA). Neither the European Union nor HADEA can be held responsible for them. Further, NEH gratefully acknowledges grant support from the German Research Foundation (grant number GRK2350/1).

## LIST of environMENTAL Consortium (sorted by partner no.)

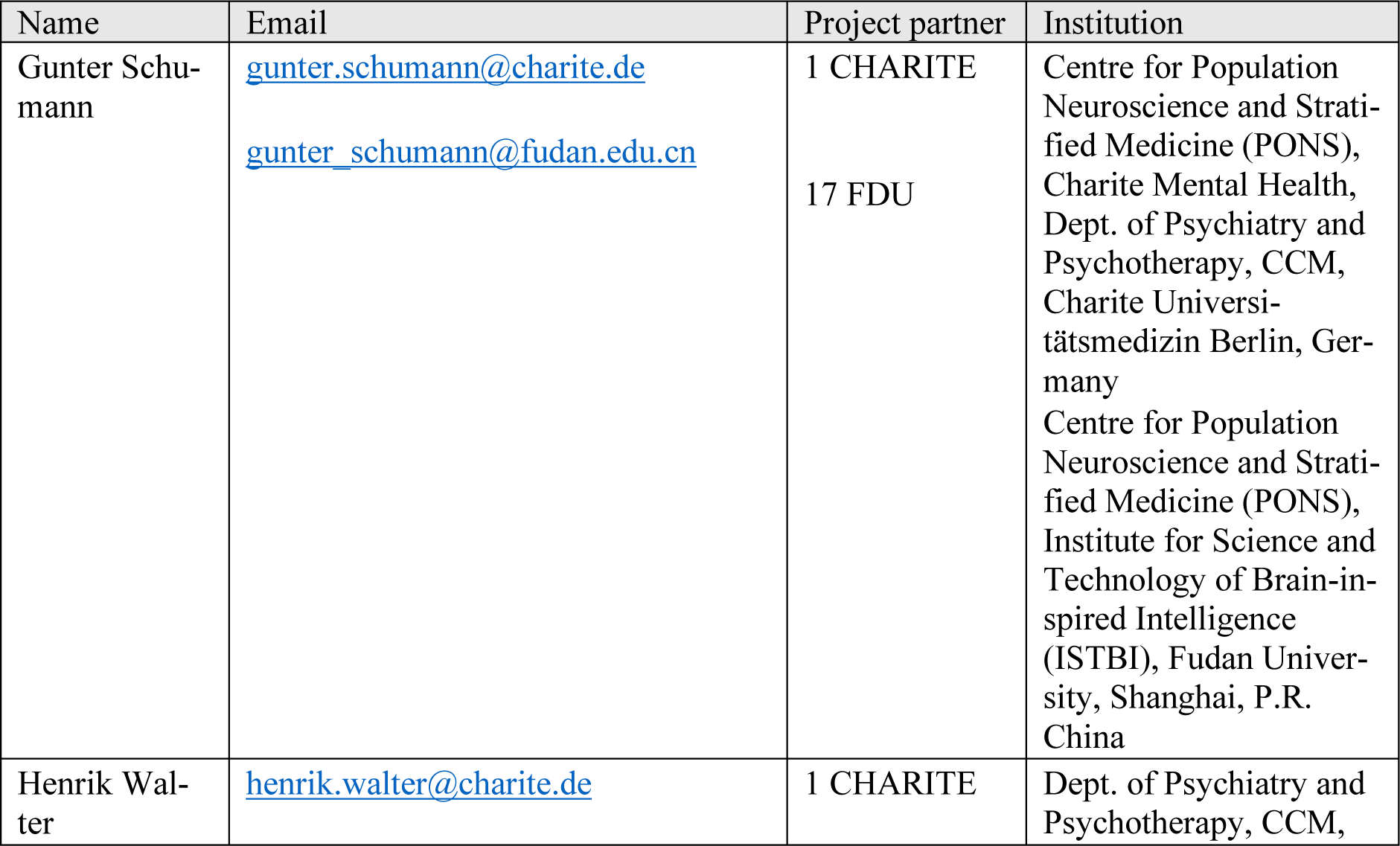
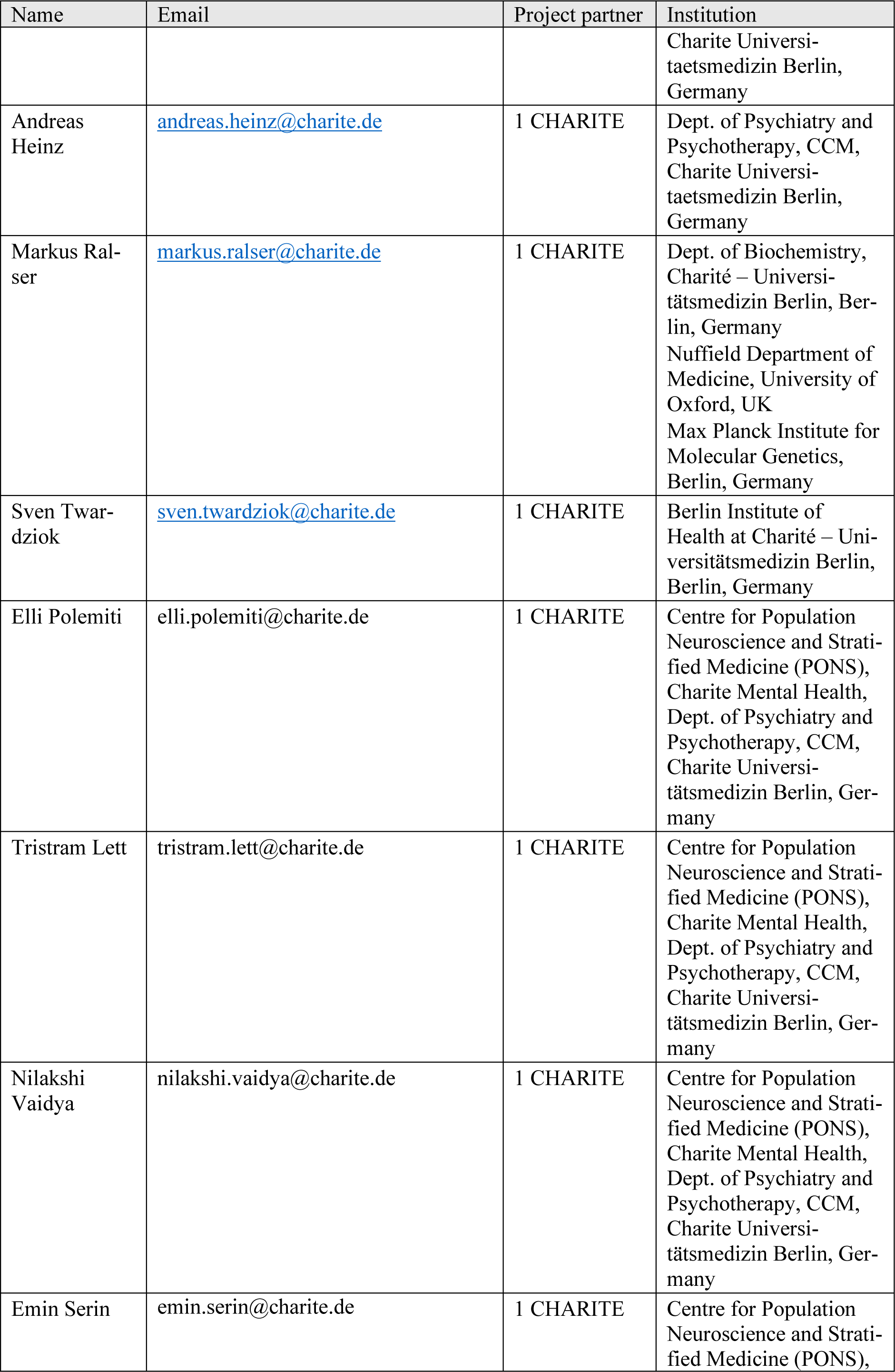
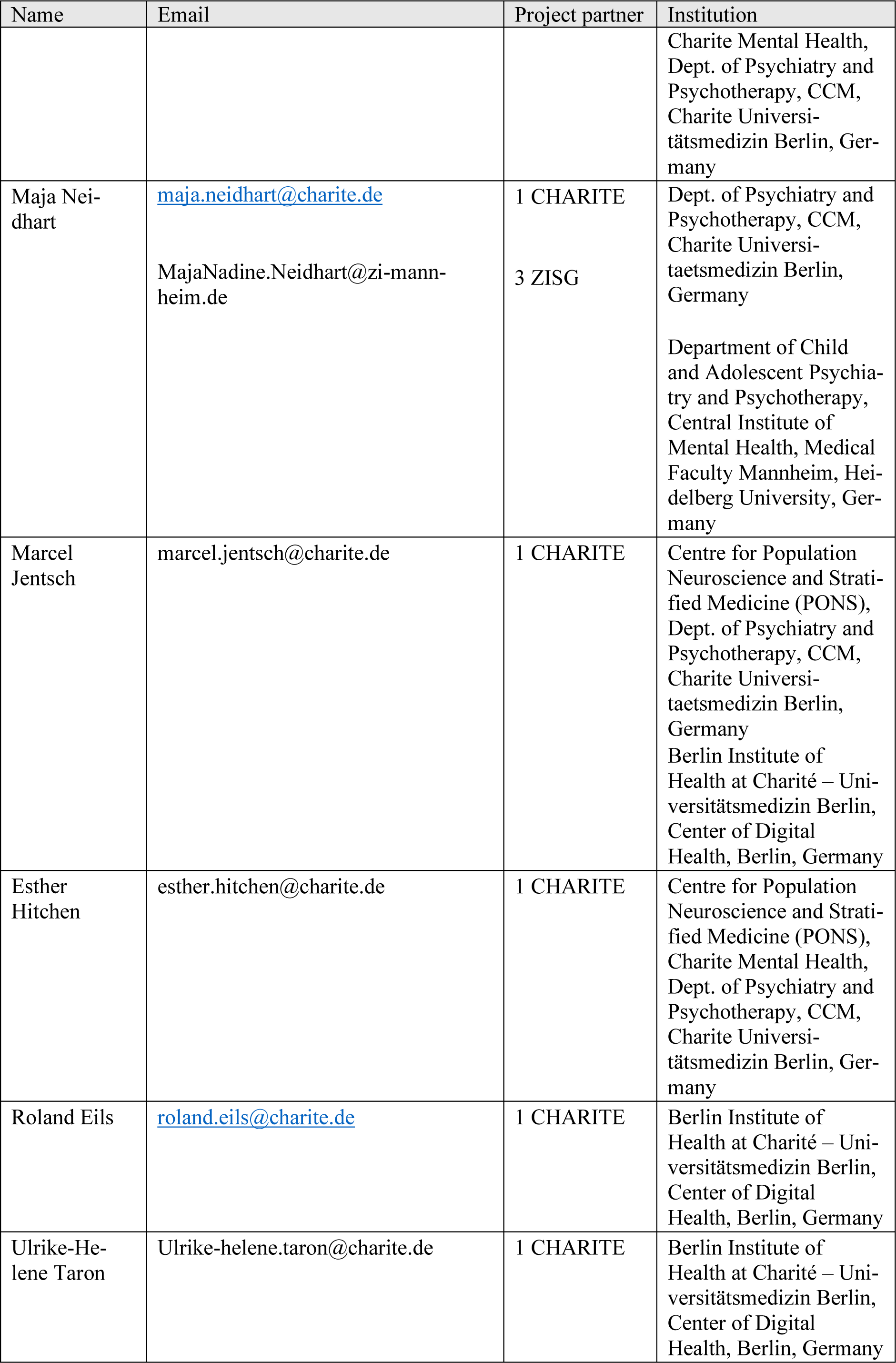
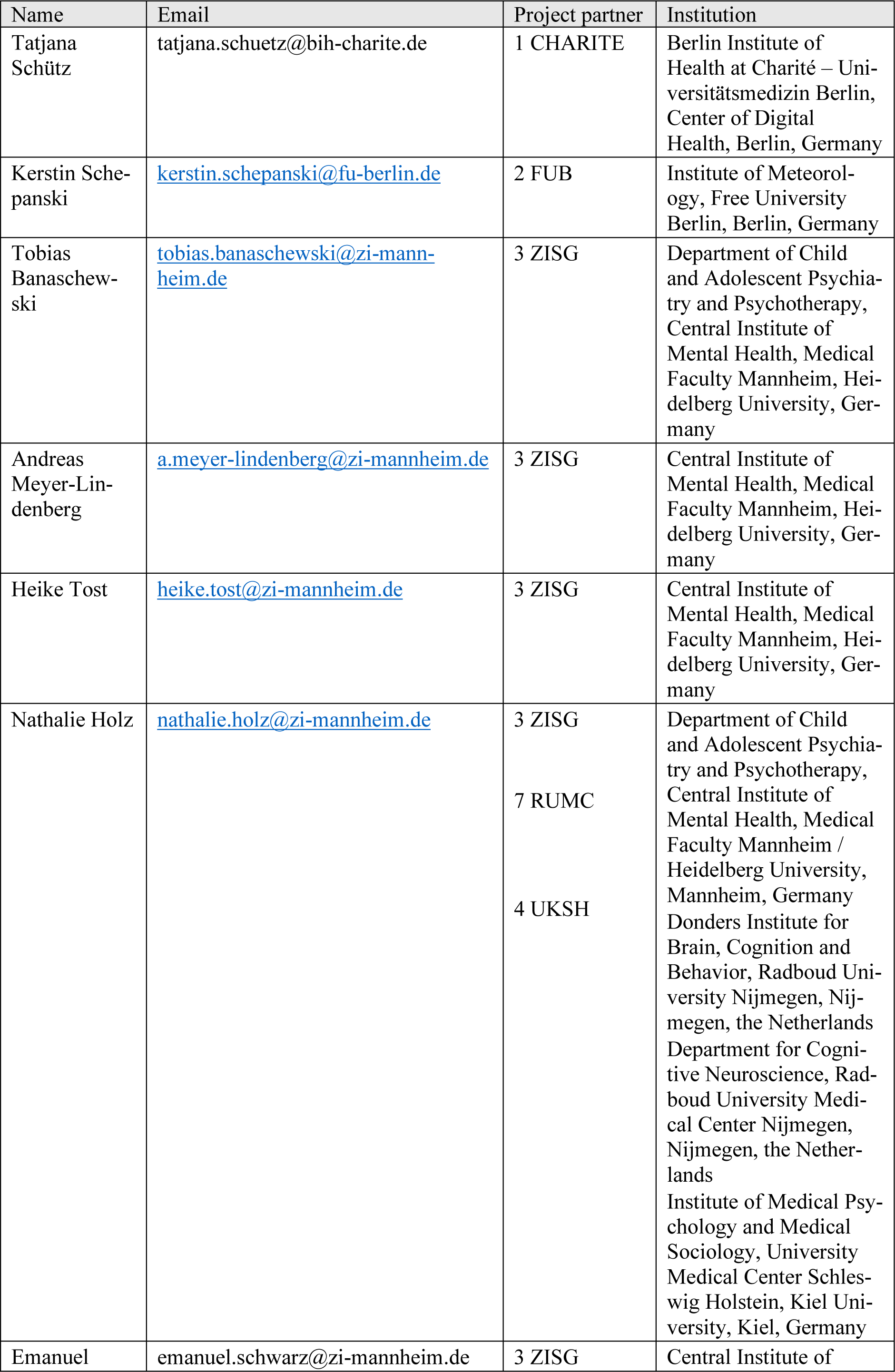
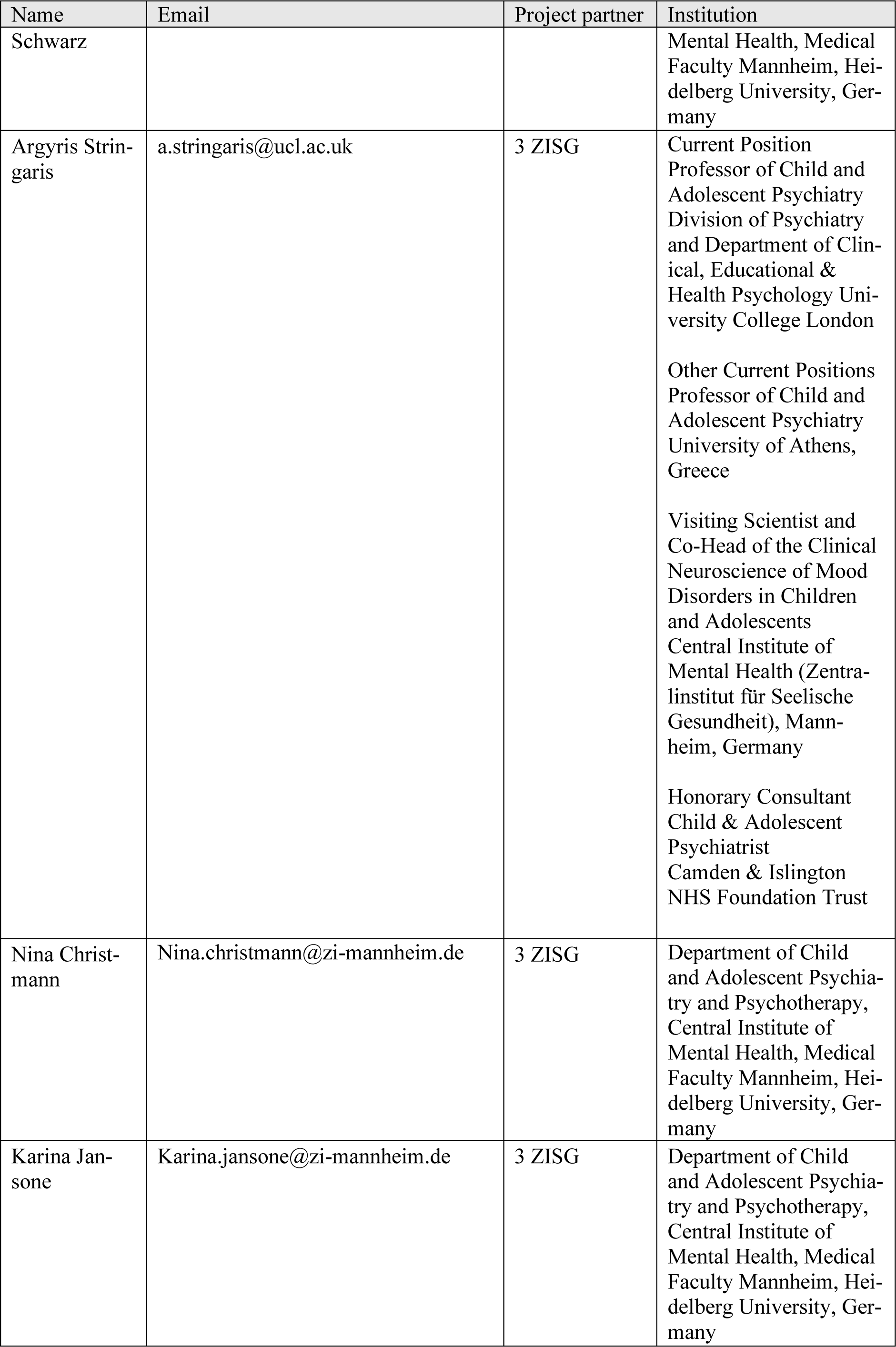
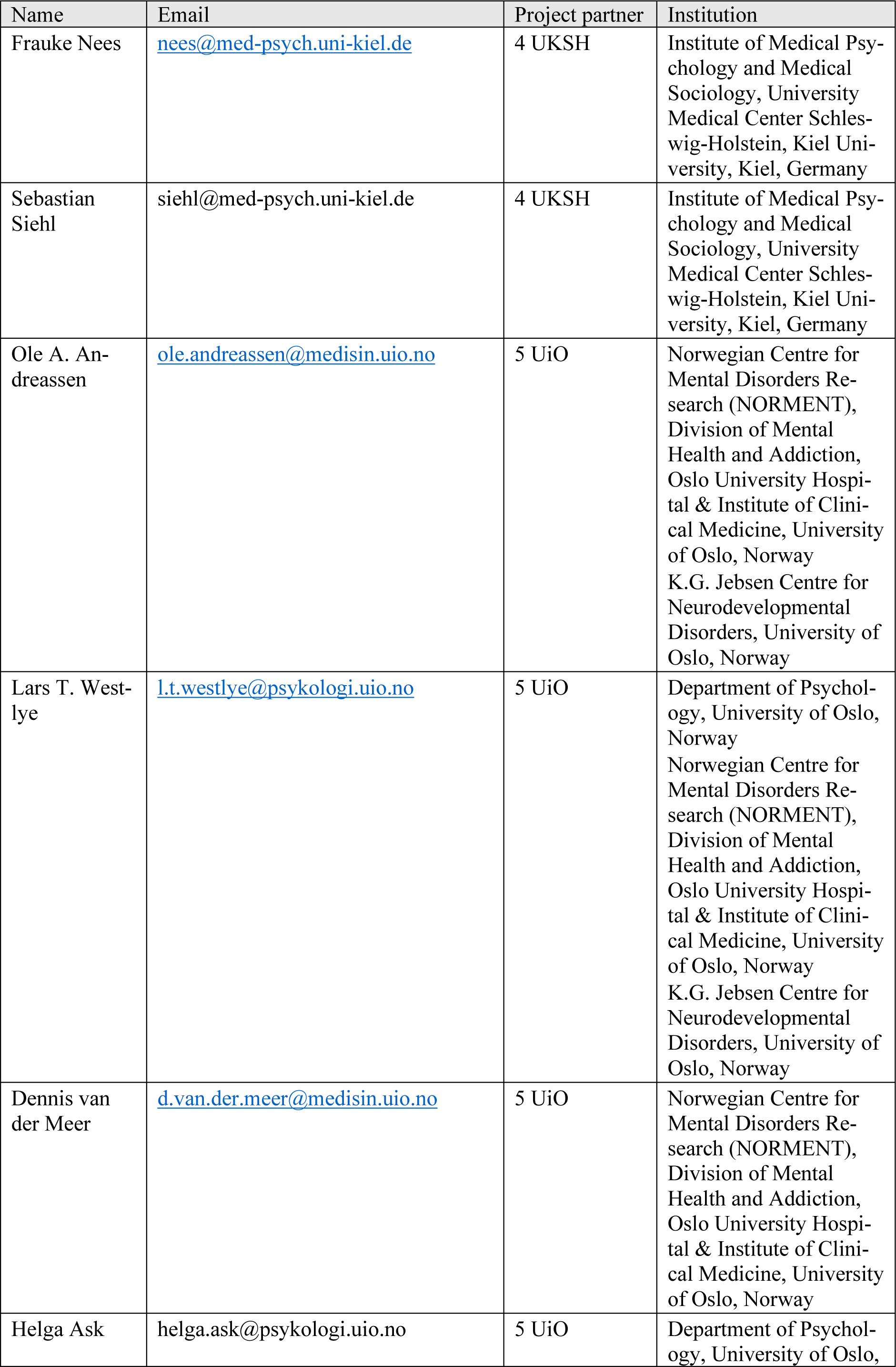
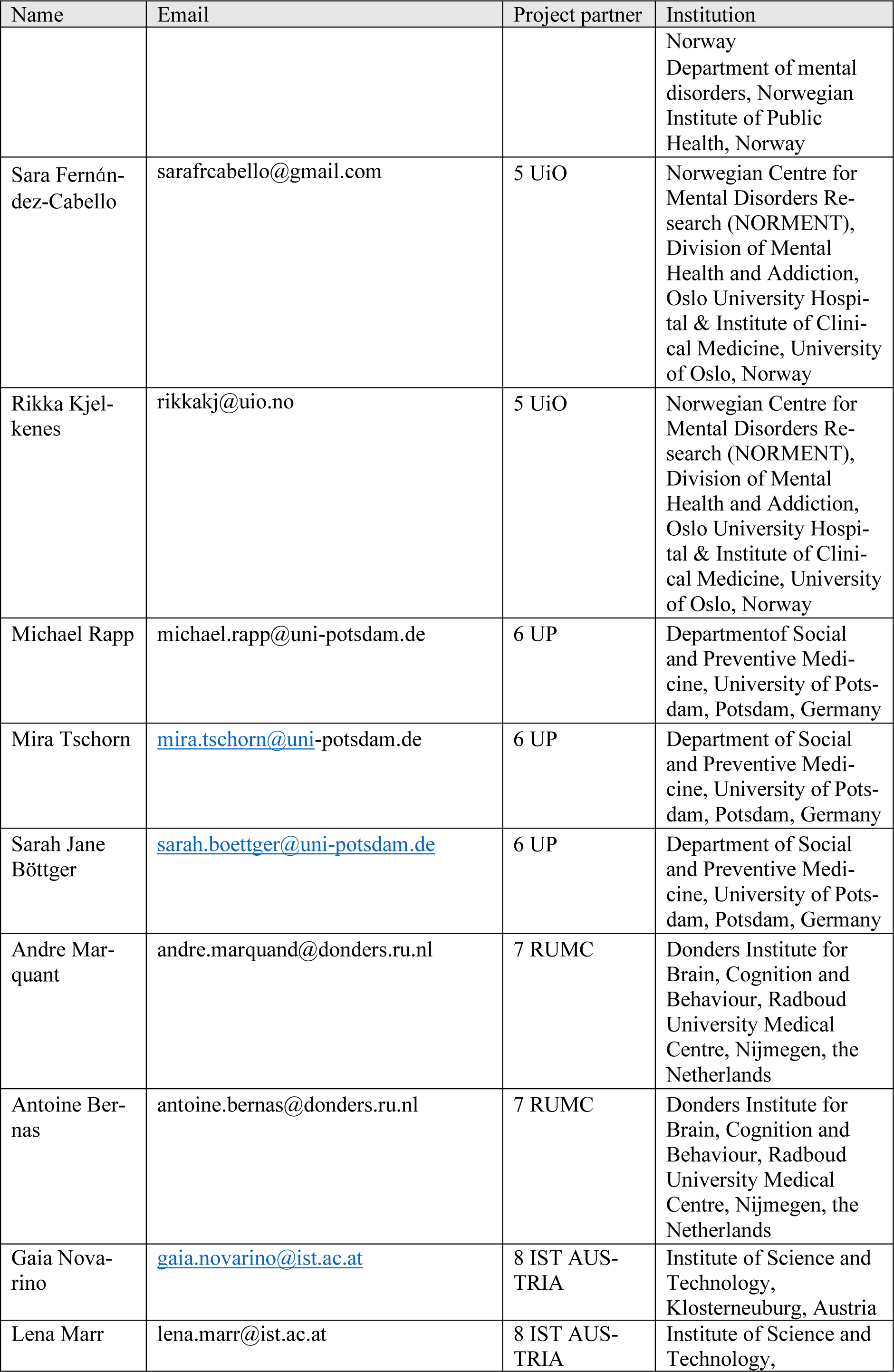
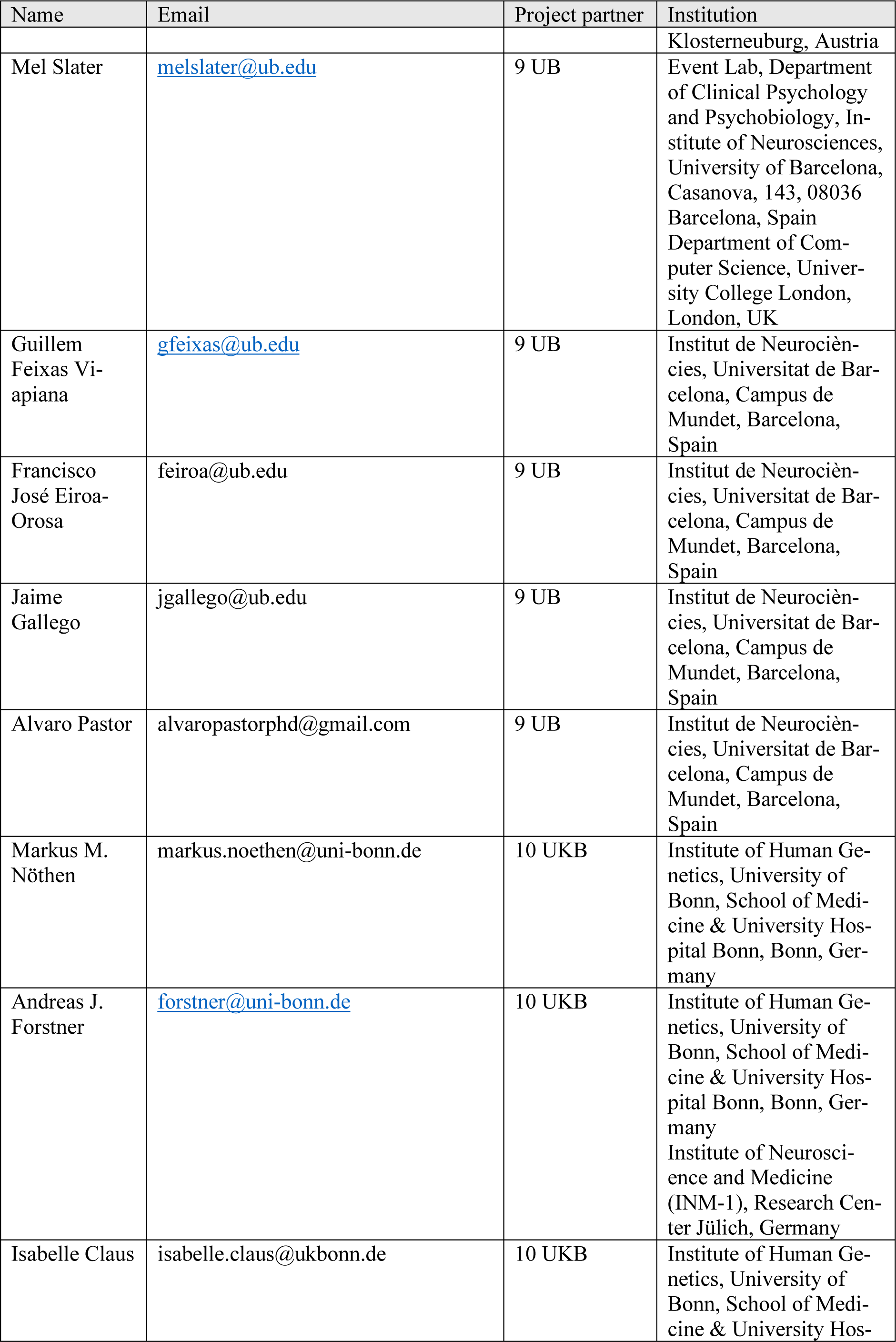
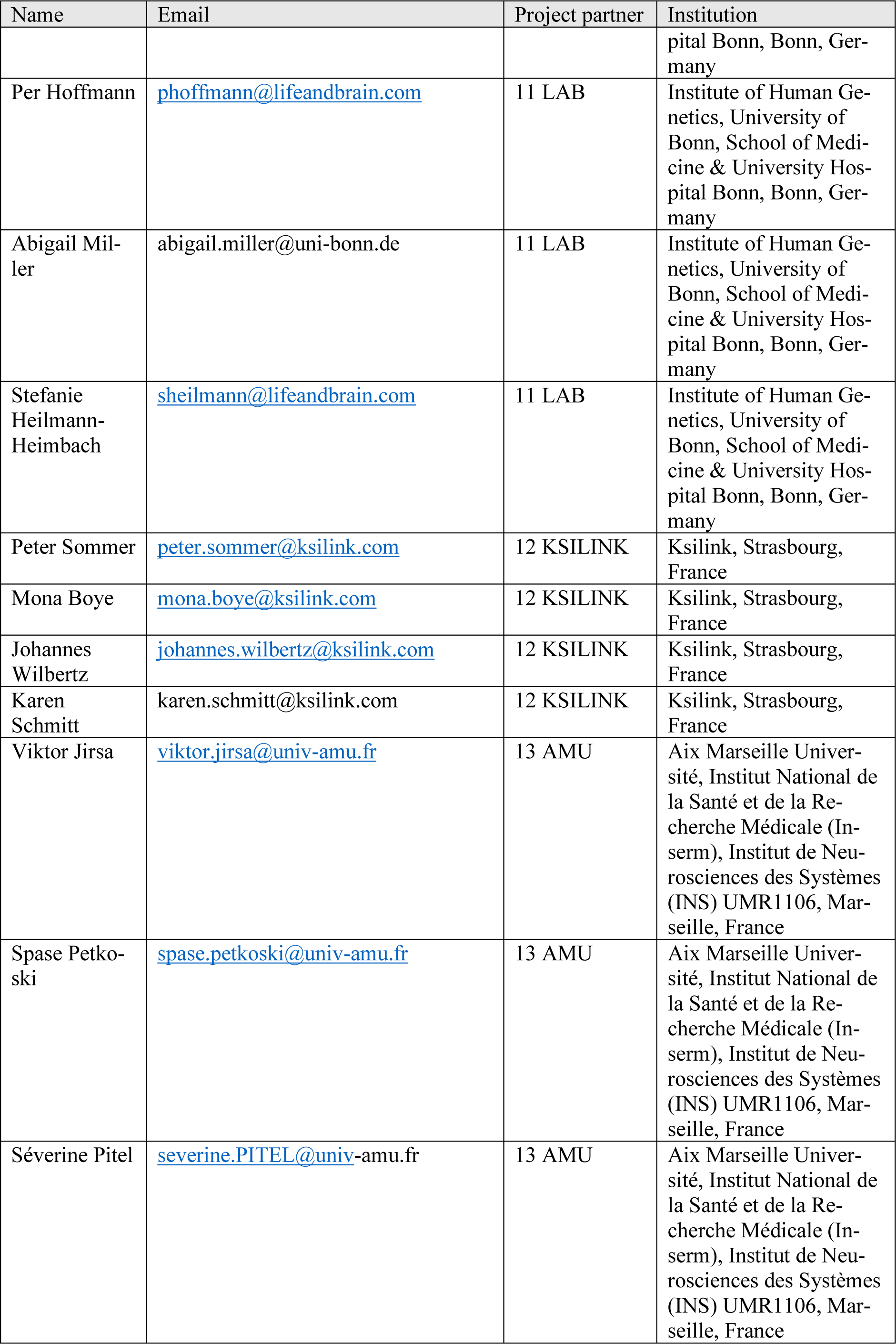
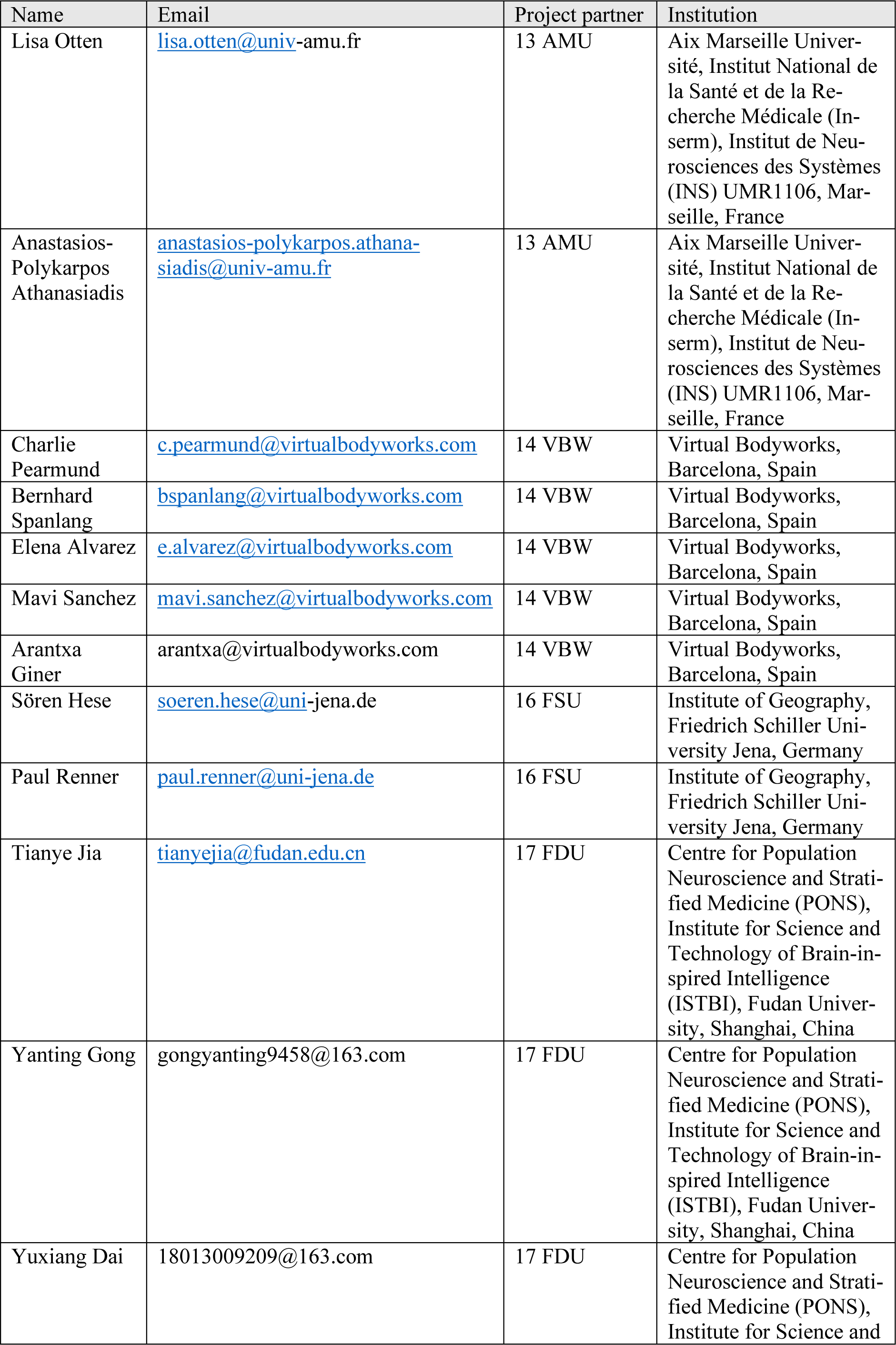
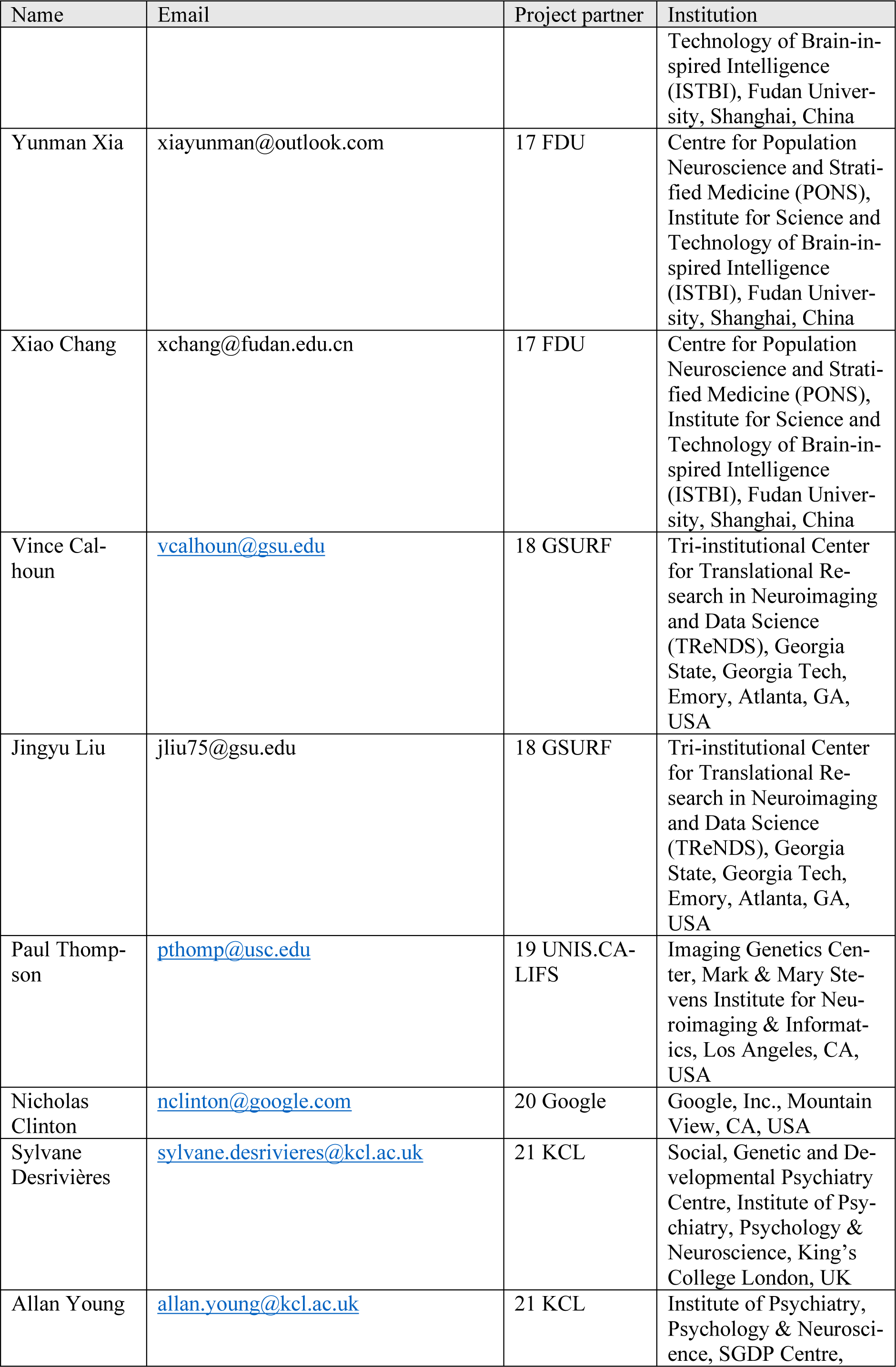
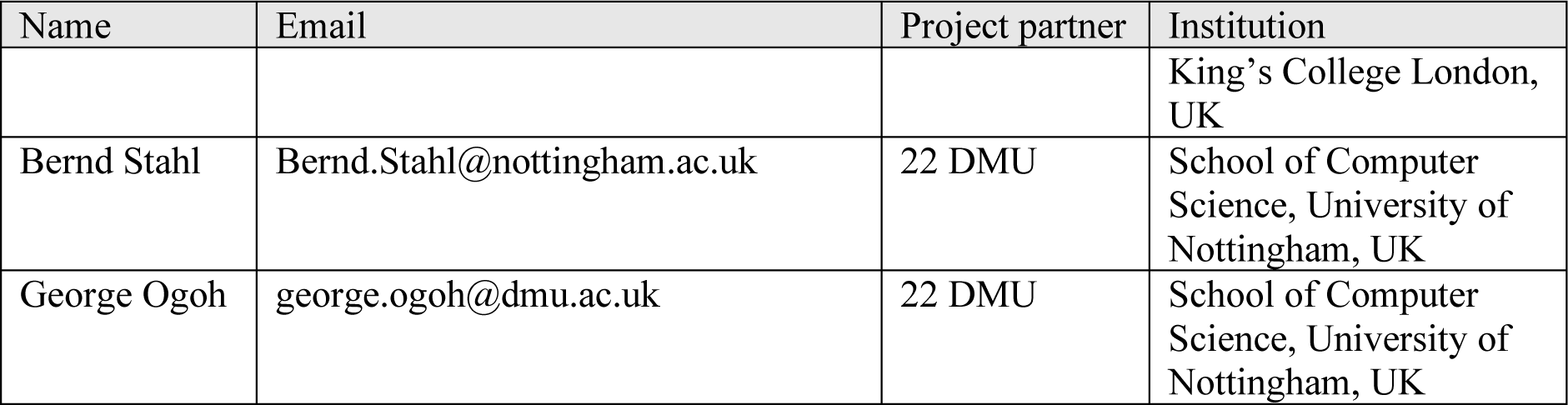

## Notes

### Competing Interest Statement

The authors have declared no competing interest.

### Funding Statement

Data used in the preparation of this article were obtained from the Adolescent Brain Cognitive De-velopment (ABCD) Study (https://abcdstudy.org), held in the NIMH Data Archive (NDA). This is a multisite, longitudinal study designed to recruit more than 10,000 children aged 9-10 and follow them over 10 years into early adulthood. The ABCD Study is supported by the National Institutes of Health Grants [U01DA041022, U01DA041028, U01DA041048, U01DA041089, U01DA041106, U01DA041117, U01DA041120, U01DA041134, U01DA041148, U01DA041156, U01DA041174, U24DA041123, U24DA041147]. A full list of supporters is available at https://abcdstudy.org/nih-collaborators. A listing of participating sites and a complete listing of the study investigators can be found at https://abcdstudy.org/principal-investigators.html. ABCD consortium investigators de-signed and implemented the study and/or provided data but did not necessarily participate in analysis or writing of this report. This manuscript reflects the views of the authors and may not reflect the opinions or views of the NIH or ABCD consortium investigators.
Additional support for this work was made possible from grants R01ES032295, R01ES031074, R01DA049238, and NSF grant 2112455, as well as the NSFC grant 82150710554 and Horizon Europe project ′environMENTAL′ 101057429. Funded by the European Union. Complementary funding was received by UK Research and Innovation (UKRI) under the UK government′s Horizon Eu-rope funding guarantee (10041392 and 10038599). Views and opinions expressed are however those of the author(s) only and do not necessarily reflect those of the European Union or European Health and Digital Executive Agency (HADEA). Neither the European Union nor HADEA can be held re-sponsible for them. Further, NEH gratefully acknowledges grant support from the German Re-search Foundation (grant number GRK2350/1).

### Summary of Updates

Author list has been updated

