## Supplementary materials for "“Urban-Satellite” estimates in the ABCD Study: Linking Neuroimaging and Mental Health to Satellite Imagery Measurements of Macro Environmental Factors"

### 2. Satellite data sources for public health research

#### *2.2. Methods for satellite data extraction and processing*

Increased satellite imagery availability, alongside advancements in analytical methods and computational platforms, has boosted access to remotely sensed derived products for health research. Different types of remotely sensed derived products exist, including derived spectral indices that can be used to depict specific characteristics of Earth's surface (e.g., built-up areas, vegetation and bodies of water), distinct types of land use / land cover and population / demographic datasets.

Remotely sensed spectral indices, calculated per pixel through mathematical operations and band combinations, aid in modeling or inferring Earth surface processes and types. Commonly used for disease mapping, risk assessment, and prediction, they capture meteorological and climatological properties, land use and land cover characteristics (vegetation, bodies of water and water content, burnt areas etc.) and urban extent [1].

Vegetation Indices (VI) such as the Normalized Difference Vegetation Index (NDVI) [2] are used for evaluation of vegetation cover and growth based on the assumption that reflectance from vegetation depends on its chemical and morphological characteristics, hence allowing to assess vegetation conditions [3].

Land Surface Temperature (LST) is used in disease predictions and public health applications, captured via satellite or airborne remote sensing by approximating Earth's emitted radiation in the Thermal Infrared channel (TIR) [4]. Unlike in-situ measurements, remotely sensed LST offers spatial and temporal availability without relying on weather stations and interpolation methods aiding in analyzing temperature impacts on mental health. LST estimation has been done using MODIS [5] and Landsat [6], with several studies investigating its impact on mental health .

##### 2.2.1. Land cover / Land use analytical products

Remotely sensed products enable researchers to use land use/land cover (LULC) classification maps for assessing human-environment interaction [10]. Satellite-derived LULC maps, created using various analytical techniques [11] are cost-effective compared to ground surveys, offering multi-temporal availability and large spatial coverage [12]. Moreover, land use/land cover change (LULCC) classification helps understand Earth's surface processes and changes [13] [14].

Specific LULC products like forest cover [15], water bodies [16] and urban environment have been developed. While initially built-up land classification maps relied on coarse resolution images, newer classification maps like Global Human Settlement Layer (GHSL) [17], Global Urban Footprint (GUF) [18], and the World Settlement Footprint (WSF) [19] leverage higher resolution imagery. Some of the LULCC products include the Global Land Analysis & Discovery (GLAD) Landsat Analysis Ready Data (ARD) – a 16-day time-series of 30m tiled Landsat normalized surface reflectance from 1997 to present, updated annually, and designed for land cover monitoring at global to local scales [20]; the Global Land Cover 2000 - a global land-cover classification product at 1 km resolution [21]; the International Geosphere-Biosphere Programme (GBP) DISCover land-cover classification product (IGBP_DISCover) (1km resolution) [22]; the Moderate Resolution Imaging Spectroradiometer (MODIS) land-cover product (MOD12Q1)(500m resolution) [23]; the global land-cover map (GlobCover) (300m resolution) [24]; the European Space Agency (ESA) Climate Change Initiative land-cover product (CCI_LC) (300m resolution) [25]; Global Land Cover - SHARE (GLC-SHARE) - a global land cover database with spatial resolution of 30 arc-seconds (~1 sq.km) [26]; and Open Street Map (OSM) Landuse/Landcover product [27].

2.2.2. Socio-economic and demographic characteristics

Besides capturing Earth's physical properties, remotely sensed products also map human activity and population distribution by combining of satellite data with socio-economic information (e.g., census data), to infer pixel-level demographic properties, exemplified by products such as the Gridded Population of the World (GPWv4.11) [28], Global Rural Urban Mapping Project (GRUMPv1) [29], LandScan Global Population Database (Landscan Global) [30], WorldPop [31], among others.

### 4. UrbanSat variables in the ABCD Study

#### *4.1. Sample description*

Address 1 was treated as the primary address where participants spent the most time. Secondary and tertiary addresses were also available if the percentage of time spent in primary address was less than 80%.

#### *4.2. Data analysis*

To unify the various data sources of the Urban-Satellite variables, we aggregate each input source into new raster files with identical extent, pixel size, and pixel locations. The aggregated rasters are built with a custom Python script employing the Geopandas and Rasterio libraries. Output pixel locations are based on a regular grid with 30 arc-second spacing (approximately 1km) covering the 48 contiguous US states from 124.8° W to 66.9° W and 24.4° N to 49.4° N. For each cell in this grid, we find every input pixel whose center lies within the cell bounds, then calculate the sum, mean, or percent value for each product as specified.

Input data for each dataset were obtained for the year 2017 to align with the baseline ABCD Study visit timing (October 2016 thru October 2018) and comprised LULC (Figure 2 and Table 2), NTL and population data (Figure 3) and spectral indices (Figure 4).

4.2.1 Land Use and Land Cover (LULC)

The land use/ land cover data from 2017,derived from the Copernicus Global Land Service [32] categorizes urban build-up, forest, crop, grass, and water areas using satellite-based discrete classifications at 100m resolution globally (Figure 2) . Urban build-up, forest, crop, grass, and water areas are identified using the Copernicus satellite-based discrete classifications (Table 2). This classification assigns a single category to each pixel at 100m resolution, globally. The forest product linked to the ABCD Study includes twelve different forest classifications. Each of the following classes: grassland, cropland, built-up area, and water corresponds to one Copernicus classification. Each aggregated raster for these categories then signifies the percent of pixels in each grid cell that belong to each classification. Figure 2 in the main paper shows a sample of these land use and land cover products at the national level.

Seasonal water data are also derived from the Copernicus Global Land Service. Unlike the classification-based data above, the source seasonal water data from Copernicus is depicted as a percentage of each 100 m input pixel. Therefore, the input 100 m pixels from Copernicus signifies both a classification belonging to one of the groups above and partial seasonal water coverage. For the seasonal water product linked to the ABCD Study, each output pixel indicates the overall percent of seasonal water coverage.

Data from the Copernicus Global Land Service are obtained as separate 20° by 20° tiles that are mosaiced into a single raster covering the 48 contiguous US states[1].

4.2.2 Nighttime Lights

[1] We note that this mosaic process results in some input pixels to shift by up to 100 m due to imperfect pixel alignment at the edges of input rasters.

The nighttime light data sourced from the Earth Observation Group (EOG) Annual VNL V2 product [33] provide average monthly radiance at about 500m resolution, aggregated to sum annual radiance values within each 1km output pixel. These data provide an average monthly radiance at an original resolution of 15 arc-seconds (approximately 500 m). The VNL 2 data are based on VIIRS satellite observations and include filtering for clouds, removal of fires, and background isolation. Our aggregated nighttime light product provides the sum of annual nighttime light radiance values within each 1 km output pixel (see Figure 3a).

##### 4.2.3 Population

Population data from 2017 are based on WorldPop Population Counts [34], specifically the US unconstrained top-down 100 m resolution dataset. These data take population census counts and use other geospatial data to disaggregate census tract information into 100 m by 100 m pixels. Our aggregated population raster sums the WorldPop populations within each 1 km pixel. Figure 3b presents the population datasets at the national and regional levels.

##### 4.2.4 Spectral Indices

We calculated three different spectral indices using 2017 Sentinel-2 Multispectral Instrument Level-1C data accessed through Google Earth Engine (GEE). These indices include the Normalized Difference Vegetation Index (NDVI), Normalized Difference Water Index (NDWI), and Normalized Difference Built-up Index (NDBI) (Figure 4).

NDVI is calculated at the level of the pixel based on the Near Infrared (NIR-Band 8A) and Red (RED-Band 4) bands of Sentinel-2 satellite image collections. As a normalized difference index, NDVI values range from -1 to 1, with higher values indicating generally denser, thicker, and greener vegetation. NDVI values depend on specific vegetation types, but in general, values below 0.2 indicate bare land, values from 0.2 to 0.5 indicate sparse vegetation, and values over 0.5 indicate dense, leafy vegetation [35]. We therefore use the threshold of 0.2 as an indicator of vegetation, and our aggregated raster indicates the percentage of each pixel with NDVI over 0.2.

We calculate the NDWI [36] to monitor surface water bodies. NDWI is calculated using the Green (Green – Band 3) and Near Infrared (NIR- Band 8A) bands of Sentinel-2 satellite image collections. NDWI values range from -1 to 1. Following [37], who finds that NDWI values over 0.3 are indicative of permanent water bodies, our aggregated product presents the percentage of each pixel with NDWI over 0.3.

We calculate NDBI from the Sentinel-2 Shortwave Infrared (SWIR-Band 11) and Near Infrared (NIR-Band 8A) bands. NDBI values range between -1 and +1. In general, the lowest NDBI values represent water bodies and vegetation, intermediate values often indicate buildings (or built up land cover), and the highest values represent open or barren areas [38,39]. As demonstrated in previous studies [40], the transition from vegetated to built-up areas often occurs with NDBI values just below 0. Figure 4 clearly demonstrates that the highest values of NDBI are present in relatively barren areas such as observed in the western US. Because built-up land cover is represented by neither the high nor low end of the NDBI spectrum, one cannot use a single threshold value to represent built-up areas as with the approach to NDBI or NDVI. Our aggregated product therefore presents the average NDBI value within each 1 km pixel.

Although NDBI does not allow for a strict threshold indicating built-up land area, NDBI values do provide details to help identify the relative concentration of built-up area. For instance, where vegetated areas and water bodies are mixed within urban areas, built-up areas are represented by relatively higher NDBI values. Figure 4 shows, as an illustration, the aggregated NDBI product alongside the percent of forest cover in and around Washington DC, USA. Within the city, NDBI tends to be relatively higher in heavily built-up areas downtown than the NDBI observed around forests, parks, and rivers.

It should be noted that the Sentinel-2 input data for these spectral indices contain some notable discontinuities at the borders between satellite passes and is the result of differences in surface and/or atmospheric conditions during individual passes of the satellite. These patterns may carry over into the NDVI, NDWI, and NDBI rasters used here. Users should remain conscious of this when comparing values between widely separated geographic locations. Furthermore, some areas of the Sentinel-2 input are also missing data in a few isolated areas scattered around the country, due to processes for masking out cloudy pixels. These areas of missing data are on the order of 100 km2 in size. These manifest as isolated no-data cells within our spectral indices products.

#### *4.3. UrbanSat characterization and association with behavioral, cognition and brain function in the ABCD Study*

The UrbanSat data in ABCD release 5.0 include 11 variables for 3 addresses collected at baseline: Address 1 (primary address) for 11226 participants, Address 2 for 1617 participants and Adress 3 for 472 participants. Each variable indicates different aspects of regional environment profiles. Through the histogram plots of these indicators associated with the primary address (Supplemental Figure 1), we demonstrated their unique data characters with remarked different shapes of distribution. In parallel, these indicators have also shown varying levels of relatedness. We present Pearson correlation across all indicators in Figure 5 (left), which lists correlations ranging from -0.84 to 0.78. The significant inter-relationship among variables is clearly visible. For instance, forest land use is most significantly negatively related to built-up land use, and most significantly positively related to NDVI, both sharing more than 50% of variance. It also relates to NDBI, nighttime lights, and population with strong correlations (absolute r>0.4). In contrast, NDWI relates to other indicators with moderate effect sizes except for permanent inland water.

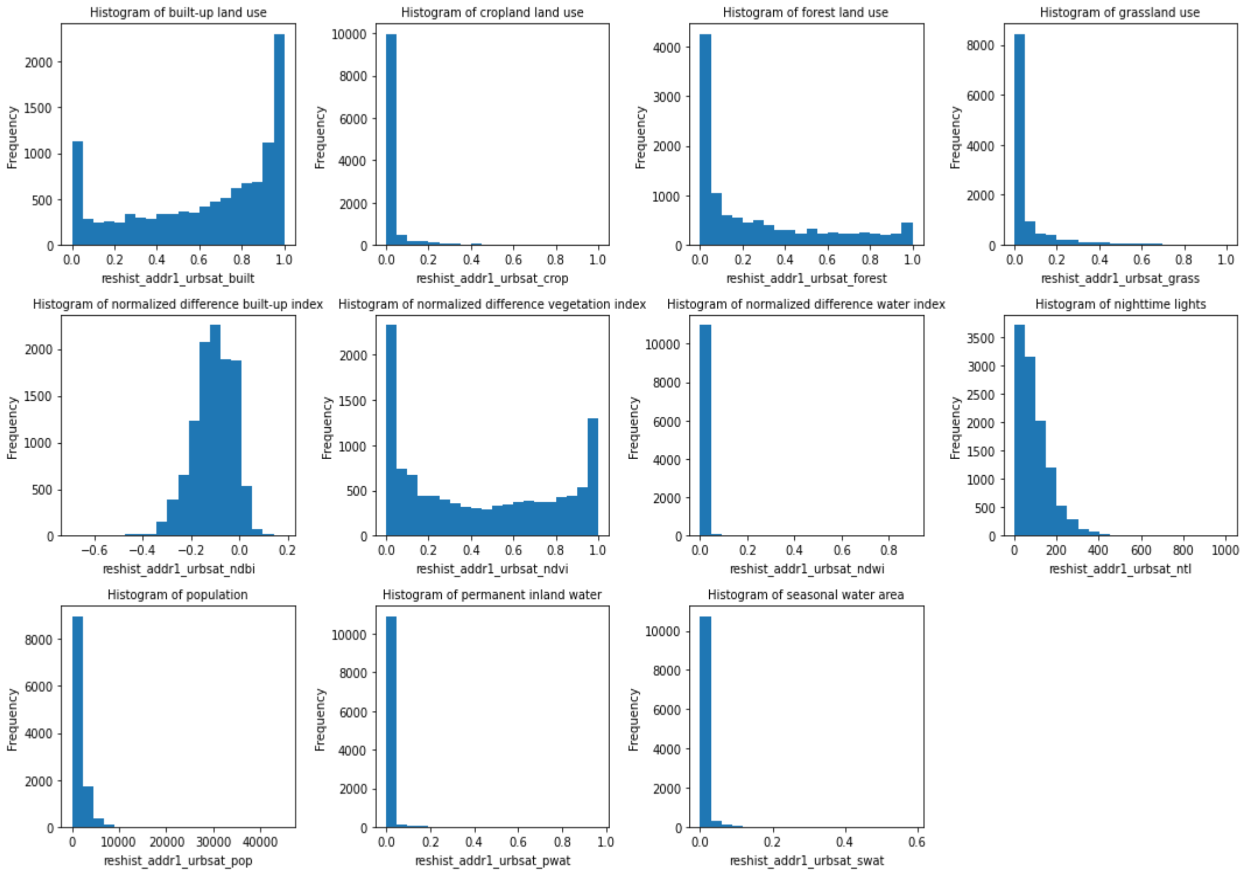


Supplemental Figure 1: Histogram of 11 UrbanSat indicators. The UrbanSat dataset consists of 11 variables from a total of 11,232 participants associated with three addresses. These indicators are built-up land use, crop land use, forest land use, normalized difference built-up index (NDBI), normalized difference vegetation index (NDVI), normalized difference water index (NDWI), nighttime lights, population, permanent inland water and seasonal water area. After removing the missing values associated with the primary address, the final number of participants was 11,006 for the histogram distribution and cross correlation of UrbanSat indicators of the primary address

4.3.1 Behavior and cognition

We selected one variable for each category from the full comprehensive assessments available in the ABCD Study from the baseline visit when the children were 9-10 years old and tested their associations with UrbanSat exposures. Child Behavior Checklist (CBCL) is a parent-rating component of the Achenbach System of Empirically Based Assessment (ASEBA) that detects behavioral and emotional problems in children and adolescents [41]. The assessment consists of 119 items describing childhood behavior corresponding to eight syndrome scales (anxious/depressed, withdrawn/depressed, somatic complaints, social problems, thought problems, attention problems, rule-breaking, aggressive behavior). The total problem count (t-score standardized, mean±STD: 45.85 ±11.34, range: 24-83) was used as the representative behavioral measure in our analyses. The NIH Toolbox® cognition battery consist of seven different tasks covering episodic memory, executive function, attention, working memory, processing speed, and language abilities [42]. These measures form three composite scores: a total score composite, a crystalized intelligence composite and a fluid Intelligence composite. We selected the total score composite (raw, uncorrected, mean±STD: 86.22 ±9.14, range: 44-117) to represent overall cognition in our analyses. We also confirmed that the T-score distribution did not show significant skew from symmetry.

4.3.2. Resting state functional MRI data

For brain function, we utilized resting state functional MRI data with satisfying quality recommended by ABCD Consortium and applied the neuromark full automated spatially constrained independent component analysis framework [43] to extract 53 robust intrinsic connectivity networks (ICNs) using the Neuromark_fmri_1.0 template. The Neuromark framework leverages an adaptive-ICA technique that automates the estimation of comparable brain markers across subjects, datasets, and studies. A set of component templates (see supplemental Table 1 and supplemental Figure 2) were used as references to guide the estimation of single-scan components for the ABCD data. These component templates were created via a unified ICA pipeline. They were constructed using the resting-state fMRI data with large samples of healthy subjects from the human connectome project (HCP) and the genomics superstruct project (GSP).

The HCP data Include 823 subjects' scans and the GSP data include 1,005 subjects' scans that passed the data QC. High model order (order = 100) group ICA was performed on each data respectively, and then the independent components (ICs) from two data were matched by examining the similarity of their spatial maps (Smith et al., 2009). The IC pairs are considered consistent and reproducible across GSP and HCP data if their spatial correlation ≥ 0.4. A correlation value ≥ 0.25 has been shown to represent a significant correspondence ($p < 0.005$, corrected) between components, and here we used a higher threshold because we would like to capture more reliable and consistent ICs as the templates. The matched IC pairs were labeled as meaningful component templates or noise components by five experts in the ICA field based on expectations that ICNs should exhibit peak activations in grey matter, low spatial overlap with known vascular, ventricular, motion, and susceptibility artifacts, and should have TCs dominated by low-frequency fluctuations inspecting the locations of the peak activations of their spatial maps and the low-frequency fluctuations of their TCs. The less noisy ICs from the GSP data were chosen as the component templates for the estimation of the single-scan components and TCs. More details of the Neuromark framework and its implementation can be found at [43].

In brief, these ICNs were organized into seven functional domains according to their anatomical locations and functional information, including subcortical, auditory, visual, sensorimotor, cognitive control, default mode, and cerebellar domains. Functional network connectivity (FNC) was computed as Pearson correlation between time courses of ICNs. Treating ICNs as nodes and FNC as weighted edges, we can represent the brain as a connected graph. In graph theory, the local clustering coefficient is a measure that quantifies the degree a node is close to its neighbors in the form of closed triplet graph. Its value ranges from 0 to 1. When a node has a high clustering coefficient, it tends to form a dense cluster with its neighbors, whereas a node with a low clustering coefficient is loosely connected to its neighbors. For a weighted graph, we implemented a clustering coefficient defined in Equation 1 proposed by Onnela, et al [44]. Focusing on default mode network (DMN), it has seven ICNs as shown in Figure 5 (right), including anterior cingulate cortex, posterior cingulate cortex, precuneus. We computed the average clustering coefficient of seven ICNs and used it as a representation of brain function in our UrbanSat association analyses.

$$Clustering\ Coefficient\ Node\_i = \frac{\sum_{jk}(W_{ij}W_{jk}W_{ki})^{1/3}}{K_i(K_i-1)};\ K_i = \sum_{j=1:53} w_{ij} \qquad \text{Equation I}$$

where $K_i$ is the degree of node I, $w_{ij}$ is the weighted edge between node i and node j.

Table S1. Peak Coordinates of Components

| **Selected Components as Regions of Interest** | **X** | **Y** | **Z** |
|---|---|---|---|
| **Subcortical Network (SC)** | | | |
| Caudate (69) | 6.5 | 10.5 | 5.5 |
| Subthalamus/hypothalamus (53) | -2.5 | -13.5 | -1.5 |
| Putamen (98) | -26.5 | 1.5 | -0.5 |
| Caudate (99) | 21.5 | 10.5 | -3.5 |

| | | | |
|---|---|---|---|
| Thalamus (45) | -12.5 | -18.5 | 11.5 |
| **Auditory Network (AUD)** | | | |
| Superior temporal gyrus ([STG], 21) | 62.5 | -22.5 | 7.5 |
| Middle temporal gyrus ([MTG], 56) | -42.5 | -6.5 | 10.5 |
| **Sensorimotor Network (SM)** | | | |
| Postcentral gyrus ([PoCG], 3) | 56.5 | -4.5 | 28.5 |
| Left postcentral gyrus ([L PoCG], 9) | -38.5 | -22.5 | 56.5 |
| Paracentral lobule ([ParaCL], 2) | 0.5 | -22.5 | 65.5 |
| Right postcentral gyrus ([R PoCG], 11) | 38.5 | -19.5 | 55.5 |
| Superior parietal lobule ([SPL], 27) | -18.5 | -43.5 | 65.5 |
| Paracentral lobule ([ParaCL], 54) | -18.5 | -9.5 | 56.5 |
| Precentral gyrus ([PreCG], 66) | -42.5 | -7.5 | 46.5 |
| Superior parietal lobule ([SPL], 80) | 20.5 | -63.5 | 58.5 |
| Postcentral gyrus ([PoCG], 72) | -47.5 | -27.5 | 43.5 |
| **Visual Network (VS)** | | | |
| Calcarine gyrus ([CalcarineG], 16) | -12.5 | -66.5 | 8.5 |
| Middle occipital gyrus ([MOG], 5) | -23.5 | -93.5 | -0.5 |
| Middle temporal gyrus ([MTG], 62) | 48.5 | -60.5 | 10.5 |
| Cuneus (15) | 15.5 | -91.5 | 22.5 |
| Right middle occipital gyrus ([R MOG], 12) | 38.5 | -73.5 | 6.5 |
| Fusiform gyrus (93) | 29.5 | -42.5 | -12.5 |
| Inferior occipital gyrus ([IOG], 20) | -36.5 | -76.5 | -4.5 |
| Lingual gyrus ([LingualG], 8) | -8.5 | -81.5 | -4.5 |
| Middle temporal gyrus ([MTG], 77) | -44.5 | -57.5 | -7.5 |
| **Cognitive-control Network (CC)** | | | |
| Inferior parietal lobule ([IPL], 68) | 45.5 | -61.5 | 43.5 |
| Insula (33) | -30.5 | 22.5 | -3.5 |
| Superior medial frontal gyrus ([SMFG], 43) | -0.5 | 50.5 | 29.5 |
| Inferior frontal gyrus ([IFG], 70) | -48.5 | 34.5 | -0.5 |
| Right inferior frontal gyrus ([R IFG], 61) | 53.5 | 22.5 | 13.5 |
| Middle frontal gyrus ([MiFG], 55) | -41.5 | 19.5 | 26.5 |
| Inferior parietal lobule ([IPL], 63) | -53.5 | -49.5 | 43.5 |
| Left inferior parietal lobue ([R IPL], 79) | 44.5 | -34.5 | 46.5 |
| Supplementary motor area ([SMA], 84) | -6.5 | 13.5 | 64.5 |
| Superior frontal gyrus ([SFG], 96) | -24.5 | 26.5 | 49.5 |
| Middle frontal gyrus ([MiFG], 88) | 30.5 | 41.5 | 28.5 |
| Hippocampus ([HiPP], 48) | 23.5 | -9.5 | -16.5 |
| Left inferior parietal lobule ([L IPL], 81) | -47.5 | 5.5 | 22.5 |
| Middle cingulate cortex ([MCC], 37) | -15.5 | 20.5 | 37.5 |
| Inferior frontal gyrus ([IFG], 67) | 39.5 | 44.5 | -0.5 |
| Middle frontal gyrus ([MiFG], 38) | -26.5 | 47.5 | 5.5 |
| Hippocampus ([HiPP], 83) | -24.5 | -36.5 | 1.5 |
| **Default-mode Network (DM)** | | | |
| Precuneus (32) | -8.5 | -66.5 | 35.5 |
| Precuneus (40) | -12.5 | -54.5 | 14.5 |
| ccc ([ACC], 23) | -2.5 | 35.5 | 2.5 |
| Posterior cingulate cortex ([PCC], 71) | -5.5 | -28.5 | 26.5 |
| Anterior cingulate cortex ([ACC], 17) | -9.5 | 46.5 | -10.5 |
| Precuneus (51) | -0.5 | -48.5 | 49.5 |
| Posterior cingulate cortex ([PCC], 94) | -2.5 | 54.5 | 31.5 |
| **Cerebellar Network (CB)** | | | |
| Cerebellum ([CB], 13) | -30.5 | -54.5 | -42.5 |
| Cerebellum ([CB], 18) | -32.5 | -79.5 | -37.5 |
| Cerebellum ([CB], 4) | 20.5 | -48.5 | -40.5 |
| Cerebellum ([CB], 7) | 30.5 | -63.5 | -40.5 |

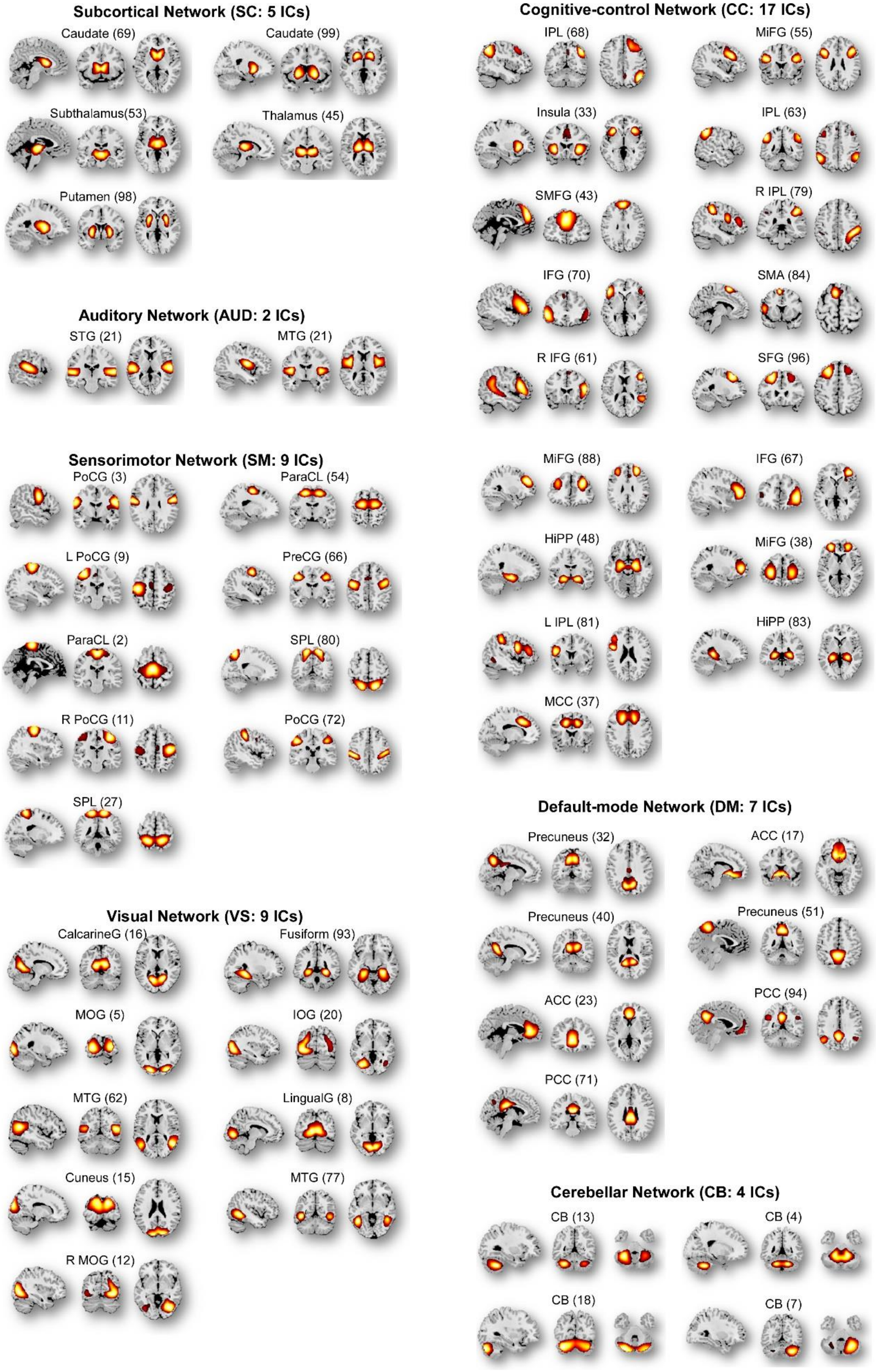
Subcortical Network (SC: 5 ICs)
Caudate (69)
Caudate (99)
Subthalamus(53)
Thalamus (45)
Putamen (98)
Auditory Network (AUD: 2 ICs)
STG (21)
MTG (21)
Sensorimotor Network (SM: 9 ICs)
PoCG (3)
ParaCL (54)
L PoCG (9)
PreCG (66)
ParaCL (2)
SPL (80)
R PoCG (11)
PoCG (72)
SPL (27)
Visual Network (VS: 9 ICs)
CalcarineG (16)
Fusiform (93)
MOG (5)
IOG (20)
MTG (62)
LingualG (8)
Cuneus (15)
MTG (77)
R MOG (12)
Cognitive-control Network (CC: 17 ICs)
IPL (68)
MiFG (55)
Insula (33)
IPL (63)
SMFG (43)
R IPL (79)
IFG (70)
SMA (84)
R IFG (61)
SFG (96)
MiFG (88)
IFG (67)
HiPP (48)
MiFG (38)
L IPL (81)
HiPP (83)
MCC (37)
Default-mode Network (DM: 7 ICs)
Precuneus (32)
ACC (17)
Precuneus (40)
Precuneus (51)
ACC (23)
PCC (94)
PCC (71)
Cerebellar Network (CB: 4 ICs)
CB (13)
CB (4)
CB (18)
CB (7)

Supplemental Figure 2. Component templates constructed by the Neuromark framework. Spatial maps of the 53 IC templates are arranged into 7 functional networks according to the anatomic and functional prior knowledge. Component templates are thresholded at |t|>10, where one-sample t-statistics have been computed across the single-subject spatial maps. Sagittal, coronal, and axial slices are shown at the maximal t-statistic for clusters larger than 3 $cm^3$.

4.3.3. Data Analysis

For the association analyses of UrbanSat indicators we utilized data from independent participants by keeping one sibling per family, to control for the impact of family structure on the results. We also filtered out data with missing values in social economic status (SES), which resulted in a dataset of 8,949 participants.

We tested the correlations between UrbanSat indicators with SES indicators (household income and parental education level). Household income is to measure the total combined family income for the past 12 months, coded as 1 to 10 from less than US $5,000 to $200,000 and greater. Parental education was coded as 0 to 21 from never attended/kindergarten only to doctoral degree. If both parents' education levels are available, the averaged value is used. Table S2 lists out the correlation results between UrbanSat indicators with education and household income using 8,716 samples.

Table S2. Correlation between UrbanSat and SES

| | Education | | Household Income | |
|---|---|---|---|---|
| | Correlation | P value | Correlation | P value |
| Build up land | -0.18 | 7.95e-71 | -0.21 | 1.46e-88 |
| Crop land | 0.06 | 1.72e-08 | 0.10 | 5.68e-21 |
| Forest land | 0.19 | 6.86e-76 | 0.18 | 1.05e-70 |
| Grass land | -0.013 | 0.22 | 0.015 | 0.15 |
| NDBI | -0.30 | 1.86e-193 | -0.28 | 1.95e-168 |
| NDVI | 0.23 | 5.45e-111 | 0.21 | 1.25e-89 |
| NDWI | 0.009 | 0.38 | 0.02 | 0.01 |
| Nighttime lights | -0.24 | 5.18e-118 | -0.30 | 6.77e-187 |
| Population | -0.20 | 2.78e-87 | -0.19 | 3.36e-77 |
| Permanent inland water | 0.02 | 0.01 | 0.017 | 0.11 |
| Seasonal inland water | 0.02 | 0.02 | 0.02 | 0.01 |

The associations between UrbanSat indicators and behavior, cognition, and brain function were tested using linear mixed effect models. The dependent variable was the CBCL total problem, the cognitive total score composite, or DMN clustering coefficient, separately. The independent predictors included UrbanSat indicators (tested individually), age, sex, with or without household income and parental education as fixed effects, and the collection site as a random effect. Bonferroni correction was applied for multiple comparison correction for 11 UrbanSat indicators.


1. Dlamini, S.N.; Beloconi, A.; Mabaso, S.; Vounatsou, P.; Impouma, B.; Fall, I.S. Review of Remotely Sensed Data Products for Disease Mapping and Epidemiology. *Remote Sens. Appl. Soc. Environ.* **2019**, *14*, 108–118, doi:10.1016/j.rsase.2019.02.005.
2. Rouse, J.W. Monitoring Vegetation Systems in the Great Plains with ERTS-1. In Proceedings of the 3rd Earth Resources Technology Satellite Symposium, 1973; 1973.
3. Huang, S.; Tang, L.; Hupy, J.P.; Wang, Y.; Shao, G. A Commentary Review on the Use of Normalized Difference Vegetation Index (NDVI) in the Era of Popular Remote Sensing. *J. For. Res.* **2021**, *32*, 1–6, doi:10.1007/s11676-020-01155-1.
4. Goldblatt, R.; Addas, A.; Crull, D.; Maghrabi, A.; Levin, G.G.; Rubinyi, S. Remotely Sensed Derived Land Surface Temperature (LST) as a Proxy for Air Temperature and Thermal Comfort at a Small Geographical Scale. *Land* **2021**, *10*, 410.
5. Neteler, M. Estimating Daily Land Surface Temperatures in Mountainous Environments by Reconstructed MODIS LST Data. *Remote Sens.* **2010**, *2*, 333–351.
6. Orimoloye, I.R.; Mazinyo, S.P.; Nel, W.; Kalumba, A.M. Spatiotemporal Monitoring of Land Surface Temperature and Estimated Radiation Using Remote Sensing: Human Health Implications for East London, South Africa. *Environ. Earth Sci.* **2018**, *77*, 77, doi:10.1007/s12665-018-7252-6.
7. Isa, N.; Naim, W.; Salleh, S. THE EFFECTS OF BUILT-UP AND GREEN AREAS ON THE LAND SURFACE TEMPERATURE OF THE KUALA LUMPUR CITY. *ISPRS - Int. Arch. Photogramm. Remote Sens. Spat. Inf. Sci.* **2017**, *XLII-4/W5*, 107–112, doi:10.5194/isprs-archives-XLII-4-W5-107-2017.
8. Widayani, P.; Yanuar, S.R.; Yogi, H.A. Relationship Analysis of Environmental Factor Change on the Evidence of Dengue Fever Diseases Using Image Transformation (Case Study: Surakarta City). *IOP Conf. Ser. Earth Environ. Sci.* **2018**, *169*, 012061, doi:10.1088/1755-1315/169/1/012061.
9. Xue, T.; Zhu, T.; Zheng, Y.; Zhang, Q. Author Correction: Declines in Mental Health Associated with Air Pollution and Temperature Variability in China. *Nat. Commun.* **2019**, *10*, 3609, doi:10.1038/s41467-019-11660-5.
10. G. Pricope, N.; L. Mapes, K.; D. Woodward, K. Remote Sensing of Human–Environment Interactions in Global Change Research: A Review of Advances, Challenges and Future Directions. *Remote Sens.* **2019**, *11*, 2783, doi:10.3390/rs11232783.
11. Qin, R.; Liu, T. A Review of Landcover Classification with Very-High Resolution Remotely Sensed Optical Images—Analysis Unit, Model Scalability and Transferability. *Remote Sens.* **2022**, *14*, 646, doi:10.3390/rs14030646.
12. Talukdar, S.; Singha, P.; Mahato, S.; Shahfahad; Pal, S.; Liou, Y.-A.; Rahman, A. Land-Use Land-Cover Classification by Machine Learning Classifiers for Satellite Observations—A Review. *Remote Sens.* **2020**, *12*, 1135, doi:10.3390/rs12071135.

13. Navin, M.S.; Agilandeeswari, L. Comprehensive Review on Land Use/Land Cover Change Classification in Remote Sensing. *J. Spectr. Imaging* **2020**, *9*.
14. Digra, M.; Dhir, R.; Sharma, N. Land Use Land Cover Classification of Remote Sensing Images Based on the Deep Learning Approaches: A Statistical Analysis and Review. *Arab. J. Geosci.* **2022**, *15*, 1003, doi:10.1007/s12517-022-10246-8.
15. Hansen, M.C.; Potapov, P.V.; Moore, R.; Hancher, M.; Turubanova, S.A.; Tyukavina, A.; Thau, D.; Stehman, S.V.; Goetz, S.J.; Loveland, T.R.; et al. High-Resolution Global Maps of 21st-Century Forest Cover Change. *Science* **2013**, *342*, 850–853, doi:10.1126/science.1244693.
16. Pekel, J.-F.; Cottam, A.; Gorelick, N.; Belward, A.S. High-Resolution Mapping of Global Surface Water and Its Long-Term Changes. *Nature* **2016**, *540*, 418–422.
17. Pesaresi, M.; Ehrlich, D.; Ferri, S.; Florczyk, A.; Freire, S.; Halkia, M.; Julea, A.; Kemper, T.; Soille, P.; Syrris, V. Operating Procedure for the Production of the Global Human Settlement Layer from Landsat Data of the Epochs 1975, 1990, 2000, and 2014. *Publ. Off. Eur. Union* **2016**, 1–62.
18. Esch, T.; Heldens, W.; Hirner, A. The Global Urban Footprint. In *Urban Remote Sensing*; CRC Press, 2018; pp. 3–14 ISBN 1-138-58664-1.
19. Marconcini, M.; Metz-Marconcini, A.; Üreyen, S.; Palacios-Lopez, D.; Hanke, W.; Bachofer, F.; Zeidler, J.; Esch, T.; Gorelick, N.; Kakarla, A.; et al. Outlining Where Humans Live, the World Settlement Footprint 2015. *Sci. Data* **2020**, *7*, 242, doi:10.1038/s41597-020-00580-5.
20. Potapov, P.; Hansen, M.C.; Kommareddy, I.; Kommareddy, A.; Turubanova, S.; Pickens, A.; Adusei, B.; Tyukavina, A.; Ying, Q. Landsat Analysis Ready Data for Global Land Cover and Land Cover Change Mapping. *Remote Sens.* **2020**, *12*, 426, doi:10.3390/rs12030426.
21. Potapov, P.; Hansen, M.C.; Pickens, A.; Hernandez-Serna, A.; Tyukavina, A.; Turubanova, S.; Zalles, V.; Li, X.; Khan, A.; Stolle, F.; et al. The Global 2000-2020 Land Cover and Land Use Change Dataset Derived From the Landsat Archive: First Results. *Front. Remote Sens.* **2022**, *3*.
22. The IGBP-DIS Global 1km Land Cover Data Set, DISCover: First Results Available online: https://pubs.er.usgs.gov/publication/70187679 (accessed on 2 December 2022).
23. Friedl, Mark; Sulla-Menashe, Damien MCD12Q1 MODIS/Terra+Aqua Land Cover Type Yearly L3 Global 500m SIN Grid V006 2019.
24. Bicheron, P.; Leroy, M.; Brockmann, C.; Krämer, U.; Miras, B.; Huc, M.; Niño, F.; Defourny, P.; Vancutsem, C.; Arino, O.; et al. Globcover: A 300 m Global Land Cover Product for 2005 Using ENVISAT MERIS Time Series. *Proceeding Second Int. Symp. Recent Adv. Quant. Remote Sens.* **2006**, 538–542.
25. Defourny, P.; Kirches, G.; Brockmann, C.; Boettcher, M.; Peters, M.; Bontemps, S.; Lamarche, C.; Schlerf, M.; Santoro, M. Land Cover CCI. *Prod. User Guide Version* **2012**, *2*, 325.
26. Latham, J.; Cumani, R.; Rosati, I.; Bloise, M. Global Land Cover Share (GLC-SHARE) Database Beta-Release Version 1.0-2014. *FAO Rome Italy* **2014**, *29*.
27. Schultz, M.; Voss, J.; Auer, M.; Carter, S.; Zipf, A. Open Land Cover from OpenStreetMap and Remote Sensing. *Int. J. Appl. Earth Obs. Geoinformation* **2017**, *63*, 206–213, doi:10.1016/j.jag.2017.07.014.
28. Doxsey-Whitfield, E.; MacManus, K.; Adamo, S.B.; Pistolesi, L.; Squires, J.; Borkovska, O.; Baptista, S.R. Taking Advantage of the Improved Availability of Census Data: A First Look at the Gridded Population of the World, Version 4. *Pap. Appl. Geogr.* **2015**, *1*, 226–234.
29. University, C. for I.E.S.I.N. (CIESIN)/Columbia; Institute (IFPRI), I.F.P.R.; Bank, W.; Tropical (CIAT), C.I. de A. *Global Rural-Urban Mapping Project, Version 1 (GRUMPv1): Population Count Grid*; NASA Socioeconomic Data and Applications Center (SEDAC) Palisades, NY, 2011;
30. Bhaduri, B.; Bright, E.; Coleman, P.; Dobson, J. LandScan. *Geoinformatics* **2002**, *5*, 34–37.
31. Tatem, A.J. WorldPop, Open Data for Spatial Demography. *Sci. Data* **2017**, *4*, 1–4.
32. Buchhorn, M.; Smets, B.; Bertels, L.; Roo, B.D.; Lesiv, M.; Tsendbazar, N.-E.; Herold, M.; Fritz, S. Copernicus Global Land Service: Land Cover 100m: Collection 3: Epoch 2017: Globe 2020.
